# Linked surveillance and genetic data uncovers programmatically relevant geographic scale of Guinea worm transmission in Chad

**DOI:** 10.1101/2020.10.05.20207324

**Authors:** Jessica V. Ribado, Nancy Li, Elizabeth Thiele, Hil Lyons, James A. Cotton, Adam Weiss, Philippe Tchindebet Ouakou, Tchonfienet Moundai, Hubert Zirimwabagabo, Sarah Anne J. Guagliardo, Guillaume Chabot-Couture, Joshua L. Proctor

## Abstract

**Background:** Guinea worm (*Dracunculus medinensis*) was detected in Chad in 2010 after a supposed ten year absence, posing a challenge to the global eradication effort. Initiation of a village-based surveillance system in 2012 revealed a substantial number of dogs infected with Guinea worm, raising questions about paratenic hosts and cross-species transmission.

**Methodology/Principal Findings:** We coupled genomic and surveillance data from 2012-2018 cases to investigate the modes of transmission between hosts and the geographic connectivity for genetically similar worm populations. Eighty-six variants across three loci on the mitochondrial genome identified 41 genetically distinct worm genotypes. Spatiotemporal modeling reveals genetically identical worms are within a median range of 18.6 kilometers of each other, but largely within approximately 50 kilometers. Genetically identical worms vary in their degree of spatial clustering, suggesting there may be different factors that favor or constrain transmission. Each worm is surrounded by five to ten genetically distinct worms within a 50 kilometer radius. In an independent population, we show that more variants revealed in whole mitochondrial genome data improved the discrimination between worm pairs.

**Conclusions/Significance:** In the largest study linking genetic and surveillance data to date of Guinea worm cases in Chad, we show genetic similarity and modeling can contribute to understanding local transmission. The overlap of genetically distinct worms in quantitatively identified transmission ranges highlights the necessity for genomic tools to link cases. The improved discrimination between worm pairs from variants identified across the complete mitochondrial genome indicates expanding genomic markers could link cases at a finer scale. These results suggest that scaling up genomic surveillance for Guinea worm may provide additional value for programmatic decision-making critical for monitoring cases and intervention efficacy to achieve elimination.

## 1. Introduction

The eradication campaign for dracunculiasis, Guinea worm disease, has made substantial progress since the first set of World Health Assembly resolutions aimed at elimination efforts passed during the International Drinking Water Supply and Sanitation Decade (1981-1990) [8]. The Guinea worm eradication campaign has decreased the global burden of disease by more than 99% [28], enabled the World Health Organization (WHO) to certify 187 countries as free from endemic Dracunculiasis transmission [3], and decreased the significant economic loss in rural settings [8]. Despite the substantial progress, endemic transmission has persisted in Angola, Chad, Ethiopia, Mali, and South Sudan [28]. In Chad, a decade long absence of reported cases was interrupted with a detected resurgence of Guinea worm cases in 2010 [45]; moreover, human cases continue to be reported [2] with a growing understanding of the role of animal reservoirs in transmission [10, 19, 38, 12, 47]. The stalled progress in Chad has led to a substantial increase in surveillance and programmatic efforts [1] as well as investments in research and novel interventions [18, 24].

Genetic sequencing is an important tool for the control of other pathogens. In the case of polio eradication, for example, these tools have been used to detect silent transmission across geographic areas and thus inform programmatic decision-making [39, 14, 23]. The inclusion of strain differentiation techniques into programmatic decision-making for poliomyletis is feasible due to the continual mutation events in the virus genome, a growing global library of samples and isolates, and a mature set of mathematical and statistical methodologies to track specific lineages. For parasites, such as Guinea worm, well-established phlyogenetic methodologies to infer high-fidelity ancestry and lineages do not yet exist [47, 15]. Nonetheless, genetic data collected from Guinea worms in Chad has already revealed research and programmatically relevant insights: human and dog hosts share a common genetic population suggesting transmission between species [47]. Genome wide data from a much smaller sample of worms confirms this finding [18]. We build upon those research insights using linked genetic and surveillance data from Chad to investigate the geographic connectivity of genetically defined worm populations.

Spatial epidemiological models have been a key tool for inferring the connectivity of different populations and providing insights into the propagation of infectious diseases. Spatial models, such as the gravity [48] and 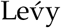 flight [7], have been utilized to describe the movement of humans and animals as a key component to the spread of infectious diseases [9, 43, 6, 52, 30, 50, 29]. These models and sophisticated inference algorithms have helped characterize the movement of pathogens and provide insights for programmatic decision-making [33, 5]. Genetic data has the potential to bypass the need for proxy human movement data by *directly* using the movement of pathogens via the genetic linkage and geographic metadata [32] even in a partially sampled transmission network [26, 17, 23]. Using genetic and epidemiological data to inform infectious disease models is a growing field, which encompasses phylodynamics [41, 53, 40, 42]. Similar to phylogenetic methodologies, phylodynamic analyses and models are most mature for virus and bacteria pathogens [31, 33, 23, 26, 17]. However, there have been recent innovations of using the biological characteristics of parasites clonally propagating in order to build phylodynamic models of malaria parasite movement within neighborhoods of Thiés, Senegal [4]. We demonstrate that probabilistic spatial models informed by Guinea worm genetic data can reveal new insights into the geographic connectivity of worm populations in Chad.

In this article, we investigate the programmatic potential of leveraging genetic data for enhanced surveillance efforts by performing a retrospective analysis and modeling of the epidemiological and genomic data collected in Chad from 2012-2018, excluding 2014. We leverage both previously reported [47] and new genetic sequences linked with surveillance data to build spatial models that reveal a consistent geographic connectedness of worm populations in Chad. We also demonstrate the additional value of expanding the number of loci sequenced for Guinea worm in order to provide higher resolution in linkages between samples. These results are followed by a discussion on the implications of increasing the scope of genetic sequencing for programmatic surveillance and decision-making.

## Materials and methods

### 1.1 DNA extraction and sequencing

Extraction was attempted on 712 worms collected in Chad from 2012-2018, excluding samples from 2014 due to availability. Whole genomic DNA was extracted from 5-15mm sections of adult female worm tissue with one of two methods: a modified Puregene DNA extraction protocol as detailed in Thiele et al. 2018 [47], or with the DNeasy Blood and Tissue Kit (Qiagen, Germantown, MD, #69582) according to the manufacturer’s protocol for tissue extraction. Five hundred and ninety-five samples were successfully sequenced for at least one locus on the mitochondria genome. Sanger sequencing and base calls were performed as detailed in [47]. There are 461 samples with all three targeted loci (CO3, cytB, and ND3-5) in our study. Thirty three of these samples overlapped with previous studies on genomic characterization in Chadian Guinea worms. [47]. The remaining 428 samples have not been published previously.

### 1.2 Alignment and variant identification

#### 1.2.1 Targeted CO3, cytB, ND3-5 loci

Sequences of targeted loci were aligned to the *Dracunculus medinensis* mitochondria genome version JN555591.1 from the European Nucleotide Archive with the BWA v0.7.17 software package [35]. Genomic ranges for each locus were determined from the start and end positions of all samples. Ranges were 3, 690-4, 308 for CO3, 2, 628-3, 234 for cytB, and 12, 562-14, 523 for ND3-5. Four hundred and sixty-one samples with successful alignment at each locus were considered for variant discovery. Alleles (A, C, G, T, or missing) were counted for each position in the identified ranges. To avoid artifacts from poor sequencing or alignment, singleton variants were recorded with their respective sample. Two samples with inflated singleton counts in cytB and ND3-5 were excluded from this study. Singleton variants identified in these two samples were excluded. Six singleton variants were identified in non-excluded samples. Remaining variants were retained if a position was missing a base call for at most one sample. Four samples had a missing position in retained variants. These criteria identified 86 variant positions concatenated to create a molecular barcode. Forty-one unique barcodes were identified across 459 worms. Barcodes were assigned an identifying number based on the number of samples belonging to each barcode.

#### 1.2.2 Untargeted complete mitochondria genome

Nineteen publicly available, untargeted mitochondria DNA sequences were downloaded from the European Nucleotide Archive [18]. Low quality bases with a minimum mapping quality of 20 were trimmed from the ends of reads. Trimmed reads were aligned to the *D. medinensis* mitochondria genome with the BWA v0.7.17 software package [35]. Variants were called on aligned reads following best practices outlined by GATK v4.1.4. Known variants are typically recommended to correct sequencing errors that lead to spurious variant calls. A set of known variants is not available for the *D. medinensis* mitochondria genome. Instead, bootstrap base recalibration was done using higher confidence calls (QD >2.0, FS >60.0, MQ <40.0, MQRankSum < −12.5, ReadPosRankSum < −8.0) until the final calls converged (2-3 steps) [49]. Final variants were filtered using the following input parameters for GATK: FS <= 13.0 or missing, SOR <3.0 or missing, and −3.1 <= ReadRankPos, BaseQRankSum, MQRankSum, ClippingRankSum <= 3.1 [18]. Given the low sample size for recalibration, orthogonal variant calling was performed using the bcftools program [34]. Genotype maximum likelihoods were determined using the mpileup command with parameters specifying a minimum mapping quality of 20 and ploidy of 1. For both variant call sets, variants found within positions 5950-6670 were removed due to high heterozygosity and 2× mapping coverage. For all other variants, a minimum depth of 10 reads was required across all samples to be included in downstream analyses.

### 1.3 Phylogenetic analysis of worms collected from humans and dogs

Phylogenetic trees were constructed using the ultrametric unweighted pair group method with arithmetic mean (UPGMA) [37]. We assumed that worms were evolving at similar rates and each base frequency is equally likely at each position. Jukes Cantor distance, the simplest substitution model meeting our assumptions, was used to calculate genetic distance between barcodes. Samples were clustered using agglomeration by average distance. Genetic distance and UPGMA clustering were calculated with R package phagorn v2.5.5 [44].

### 1.4 Linking genomic samples to epidemological data

We linked the sequenced samples with corresponding surveillance data using national case reports and standardized surveys, described in detail in [25]. Case data was collected from an active and passive village-based surveillance system by the Chad Guinea Worm Eradication Program (CGWEP) and the Ministry of Public Health (MOHP) [21]. Summary case reports at the village level are compiled across surveillance sites by the CGWEP program. Information on infected dogs was collected through a standardized survey with the self-identified owner. Where available, global position system (GPS) coordinates from the standardized surveys were validated with village coordinates from the national case reports. The GPS coordinates were consistent across national case reports and standardized surveys for 207 samples. In instances where coordinates differed between the standardized surveys and the national case report, the national case report coordinates were assigned (181 samples). For cases reported with a village name in the standardized survey but without GPS coordinates, the GPS coordinates associated with the village name in the national case report was assigned to that sample (2 samples). For cases reported with a village name and GPS coordinates, but the village name was not in the national case report, standardized survey case coordinates was assigned to that sample (37 samples). Four hundred and twenty-six worms were linked across 245 hosts. Sixty-five of the 245 hosts were infected by more than one worm. Multi-infected hosts had an median of four parasites, however, up to 24 parasites in a single host were observed.

### 1.5 Probabilistic spatial models of Guinea worm connectivity

#### 1.5.1 Spatial models

Spatial movement models have demonstrated that distance is an important factor influencing the spread of pathogens. [52, 43, 6, 26]. Here, due to the yearly transmission cycle of the Guinea worm parasite [8], we specify a spatial model similar to a probabilistic diffusion model for a discrete spatial network representing the villages affected by Guinea worm in Chad. The model describes the probability of a worm being transmitted from village *i* to village *j* based on pairwise genetic similarities *f* and distance between pairs *d* such that *p*_*ij*_ = *C*_*i*_f(*d*_*ij*_) where *C*_*i*_ is a scaling factor and f is a data-driven, empirical form directly computed from the Guinea worm data from Chad. Similar to other parametric formulations such as the diffusion, gravity [6], or Lévy flight models [26], our non-parametric formulation could be used to predict where a future linked Guinea worm case might appear.

#### 1.5.2 Pairwise genetic similarity and distance between samples

To infer the functional form of *f* in the model above, we utilize the genetic similarity between samples and the associated GPS coordinates for villages. The genetic similarity is calculated for each unique worm pair as 1 - (number of discordant bases between barcodes/total number of bases compared); this similarity metric accounts for samples having a different number of resolved bases. Discordant bases were equally penalized (−1) regardless of the base. Without a known organismal mutation rate, we assume a similar mutation rate as the *Caenorhabditis elegans* spontaneous mitochondrial mutation rate estimated at 1.05*e* − 7 site/generation [16]. From this rate, we expect 0.0015 mutations for the 14,628 base pair *Dracunculus medinensis* mitochondrial genome between generations. We have 86 variant sites across 459 worms, suggesting an average of 0.187 mutations per sample in the population. Both the average mutation per sample and the extrapolated mutation per generation from the *C. elegans* rate, are less than one, indicating a single base pair is sufficient to distinguish barcode sets. For computing the geographic distance between each pair of samples *d*, we used the haversine distance as implemented in the R package geosphere v1.5 [27].

Partitioning samples according to genetic similarity requires caution. Even grouping samples that are different by one base pair, either from a real mutation or sequencing error, creates an algorithmic issue. For example, if a genetic sequence that is one base pair different from two other sequences, but the discordance is at different base pair positions, then this three sample group would not be a single base pair different for all pairwise comparisons. In this work, we enforce that the groupings maintain the required base-pair differences by only including the pairwise comparisons that satisfy the requirement; note that this is different than grouping all of the samples with a base-pair discordance and then performing all of the pairwise comparisons. We use these relaxed similarity thresholds to identify shared identity between barcode pairs to investigate the impact of geographic clustering (§1.5.4).

#### 1.5.3 Spatial models by molecular barcode sets

To characterize the functional form *f*, we partition the samples according to their genetic similarity. Samples that are identical are grouped into molecular barcodes; for each barcode *k*, all of the pairwise distances within the set are used to compute a barcode functional form *f*_*k*_. The set of functions *f*_*k*_ are then averaged to produce a single *f*. We also investigated the impact of partitioning samples with a less restrictive criteria based on the assumed mutation rate as described above. Similar to research in the comparison of power-law distributions [11], we utilize the empirical cumulative densities *f* using stat ecdf and compare distributions using the Kolmogorov-Smirnov test with base R v3.6.3 [46]. We plot the empirical distributions as smoothed kernel density distributions using the function stat density contained in the R package ggplot2 v3.3.0 [51].

#### 1.5.4 Sensitivity analyses

We investigated the results by performing a series of sensitivity analyses including statistical bootstrapping. Distance permutations between worms pairs included 100 iterations of sampling without replacement for the distance vector. Multi-infected host bootstrap subsampling included 100 iterations of choosing a single case worm at random for each host. In instances where only one case is observed per host, that case was consistently the representative. Each iteration was considered an independent distribution and analyzed as described above. In addition, we also tested the impact of relaxed similarity thresholds for defining matched identity of the molecular barcodes between pairs on the geographic clustering.

#### 1.5.5 Analyzing the diversity of genetic samples by geography

Population worm cases (425) were ordered by ascending distance in reference to a single worm case, herein referred to as the index worm for each sample comparison. Cumulative barcode diversity scores were calculated for each worm in the study as the index case. Each worm case barcode that did not match the index case augmented the barcode diversity by one. When two worms were equidistant from the index case, the highest cumulative score was used as the barcode diversity for that distance metric. We also investigated the effect of opportunistic sampling on barcode geographic clustering by comparing the cumulative distance from an index case to every other worm in the population. From each index case, we expect the number of samples to increase at variable rates depending on sample clusters. Deviations from an increase (a sustained flat line) between samples would indicate geographic breaks between sample clusters.

### 1.6 Analyzing sensitivity of additional loci outside of CO3, cytB, and ND3-5

Variants were considered to be within loci if found within the gene ranges provided by PlasmoDB for each gene; CO3 (3, 788-4, 534), cytB (2, 628-3, 345), and ND3-5 (12, 562-14, 566). Note, these mitochondrial coordinates slightly differed from the ranges identified by target loci sequencing. Pairwise genetic similarity was calculated from barcodes created with all variants, variants within loci, or variants outside of loci as described in the previous section. For bootstrap subsampling tests, variants outside of loci ranges were randomly selected to contain the same number of variants found within loci to create a new barcode for genetic similarity comparisons. Genetic similarity tests for the different regions were robustly tested across two variant calling pipelines (GATK and bcftools mpileup, see DNA alignment and variant calling) with all variants and singletons excluded. Smooth kernel density distributions were generated with ggplot2 v3.3.0 option “stat density”. A two sided Kolmogorov-Smirnov test was used to compare genetic similarity distributions of different loci classes with base R v3.6.3 [46].

## Results

### 1.7 Genetic and epidemiological characteristics of Guinea worm cases

Four hundred and fifty-nine worms (30 humans hosts, 429 dog hosts) collected from 2012-2018 were successfully sequenced (Fig 1A). Four hundred and twenty-six of successfully sequenced worms were matched to case coordinates. Worm cases mainly clustered along the Chari River (Fig 1A). Identical mitochondrial sequences were observed for cases across years (Fig 1B). Eighty six variants were identified in the CO3, cytB, and ND3-5 loci of 618, 606, and 1, 961 base pairs, respectively. The concatenation of the 86 variants resulted in 41 unique molecular barcodes (S1 Table). Barcode accumulation curves suggest the 41 barcodes in this study are not saturating the potential diversity of the unobserved population given the proportion of low frequency barcodes in the observed population. We studied the discovery of new barcodes beyond the 41 observed with extrapolations up to 5000 samples using a standard negative binomial and a statistical, empirical Bayes approach [22]. The negative binomial procedure suggests we may observe 40 additional barcodes and the empirical Bayes approach suggests we may observe up to 15 additional barcodes for a sampled population size of 5000. Despite the differences between methods, both indicate that modestly increasing the number of samples sequenced is not likely to capture the complete underlying genetic diversity. Moreover, both methods indicate extrapolating to an even larger number of sequenced samples, 5000 or larger, may not be enough due to the abundance of low-frequency barcodes in the sampled population. However, the extrapolation is hindered by the available data; see S1 Appendix for further methodological details and numerical investigations.

**Figure 1:**
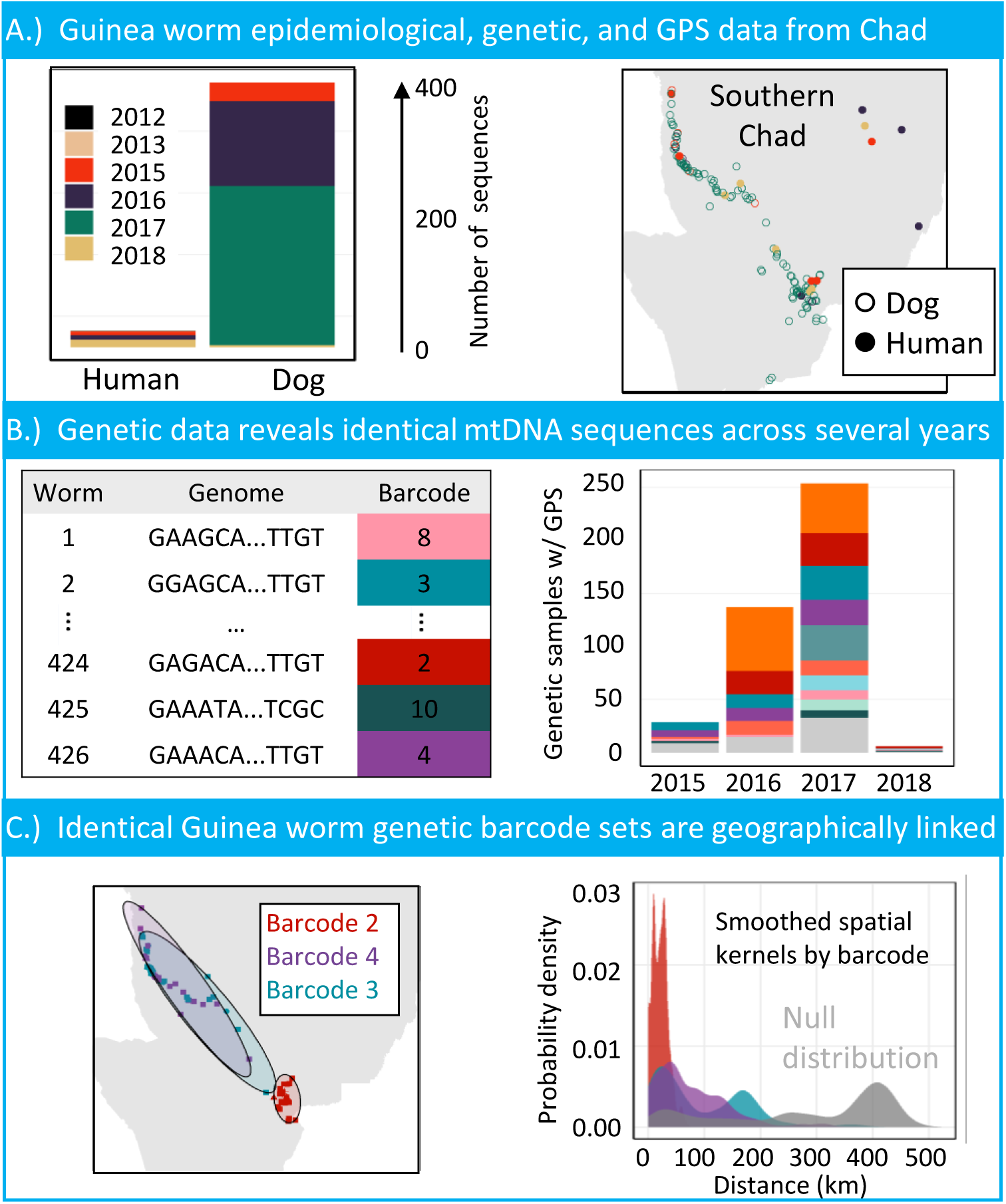
Characteristics of genetically characterized Guinea worm cases in Chad from 2012-2018. A.) Number of cases per host species collected from 2012-2018, with GPS matched sample distributions across the Chari River. B.) The number of GPS matched samples that belong to barcode sets. Note, not all barcode colors are show in the left figure. Barcodes with less than 10 samples in the population are colored in light grey for visual clarity. C.) Samples belonging to barcode sets 2, 3 and 4 are shown in their respective ranges. Smoothed kernel densities for the three barcode sets show the mass of the distributions correlate with the spatial connectedness of samples.

### 1.8 Phylogenetic similarity between host species

There is not a consistent trend of interspecies transmission for barcodes shared across host species. Phylogenetic analysis of all available Guinea worm barcodes collected from humans and dogs identifies clades of repeated or highly similar barcodes. Twelve of the 41 identified barcodes are shared between human and dog hosts. There is no clear signal of unilateral transmission between host species (S1 Fig). Barcodes were equally likely to appear in both species at the same time (barcodes 2, 6, 8, 10, 36), dogs a year prior to humans (Barcodes 5, 7, 21), or in humans a year prior to dogs (Barcodes 3, 4, 13, 14). There are more unique dog barcodes driven by the high number of reported cases in dogs (Fig 1A); however, barcodes found exclusively in one species do not form separate clades. The average nucleotide similarity (*π*) between human and dog barcodes ranged from 0.6-1.0, similar to within host nucleotide similarity ranges. Human samples are the sole genetic representative for 2012, 2013, and 2018 cases with 1, 2, and 11 samples. Only four barcodes from this set (7, 21, 40 and 41) were observed as a singular event.

### 1.9 Genetically identical worms are geographically clustered

#### 1.9.1 Population level comparisons

Pairwise genetic similarity scores for all worms with GPS data available reveals genetically identical barcodes are spatially clustered in Chad. Identical barcodes are within a median 18.6 kilometer range (standard deviation = 82.5 kilometers), and often within an approximately 50 kilometer radius (Fig 2A). Non-identical barcodes were more evenly distributed within a median 222.2 kilometers (standard deviation = 157.5 kilometers), however spread across a 400 kilometer radius, with slight inclination to be clustered within 100 kilometers or 400 kilometers (Fig 2A). The spatial distributions between identical and non-identical barcode distributions are statistically different (Kolmogorov–Smirnov test, p < 0.001). Reclassifying worm pairs with a single nucleotide difference to account for potential elevated mutation rates or a sequencing mutation as identical robustly replicated geographical clustering within a 50 kilometer radius (S2 Fig). Permuting the distance between worm pairs removed spatial clustering for identical worms (S3 Fig).

**Figure 2:**
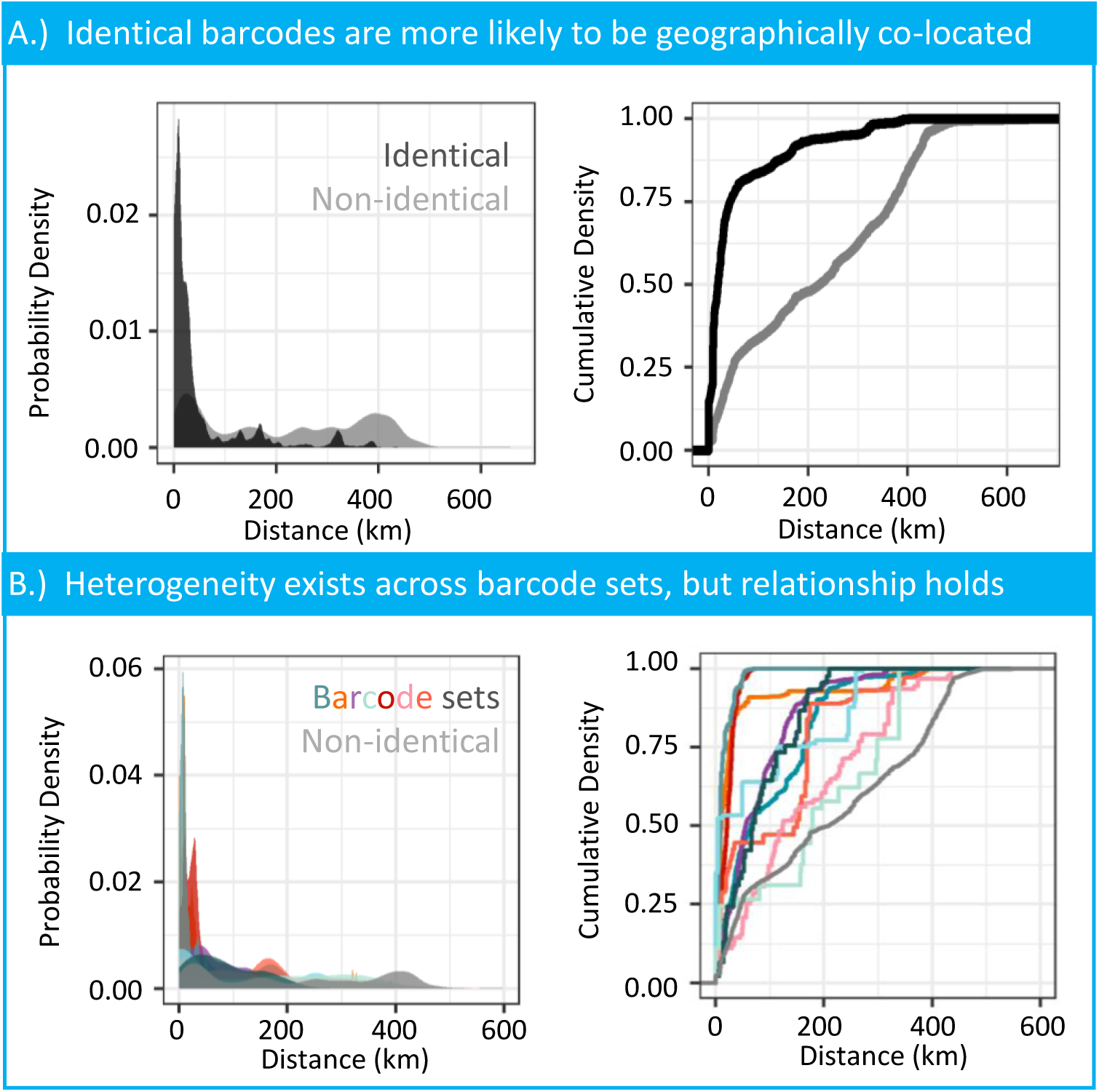
Distance density estimates of pairwise similarity for identical and non-identical barcodes. A.) Pairwise comparisons of identical barcodes in black (n=10, 695) to a null of non-identical barcodes in grey (n=79, 830). This distrubtion at 400 kilometers is the max distance between sampled cases along the Chari River. This enrichment of pirwise sample distance around 400 kilometers should not be considered the transmission upperbound. B.) The number of comparisons for each barcode are the unique pairs of all worms sharing that barcode identifier. The null distribution in grey of non-identical barcodes is subset to only include comparisons where one of the common barcode must be found in the pair (n = 56, 913).

Geographic clustering of barcodes is not due solely to the similarity of parasites within hosts that carry multiple infections. Sixty-five of the 245 hosts had multiple worm infections, with a median of 4 worms (range = 2-24) per multi-infected host (S4 Fig). Worms captured from the same host were not always genetically identical. The distribution of genetic similarity for worms with the same host did not differ from worms with the same GPS coordinates from different hosts (S4 Fig). Bootstrap subsampling of a single worm from each host maintained spatial clustering with low variability (S5 Fig).

#### 1.9.2 Individual barcode comparisons

Individual barcodes show variation in their geographical range (Fig 2B). For example in Fig 1C, cases with Barcode 2 are found in the southeast mouth of the Chari River, while cases with Barcode 3 and 4 are spread along the entire river. The geographic range for samples in each common barcode can be seen in S6 Fig. We replicated the smooth kernel density estimates for 10 of the 38 barcodes; note that three of the barcodes do not contain associated GPS locations for any sample. The geographic variability is not dependent on the total number of worms assigned to a barcode (S1 Table). Two of the barcodes seen in Fig 2B represented with pink and mint are more similar to the null distribution and found in a broader geographic in Chad.

The variation in geographical ranges is not driven by a geographical isolation of cases sharing barcodes. For most worms, we observe a steep increase in unique barcodes up to 200 kilometers (Fig 3). Worms with a slow increase in the unique barcodes after 200 kilometers correspond to the human cases observed furthest east in Fig 1A. These index worms follow a similar steep increase once the radius includes samples along the Chari River. The plateau of diversity observed in most worms around 18-20 unique barcodes is an artifact of opportunistic sampling proximity (S7 Fig).

**Figure 3:**
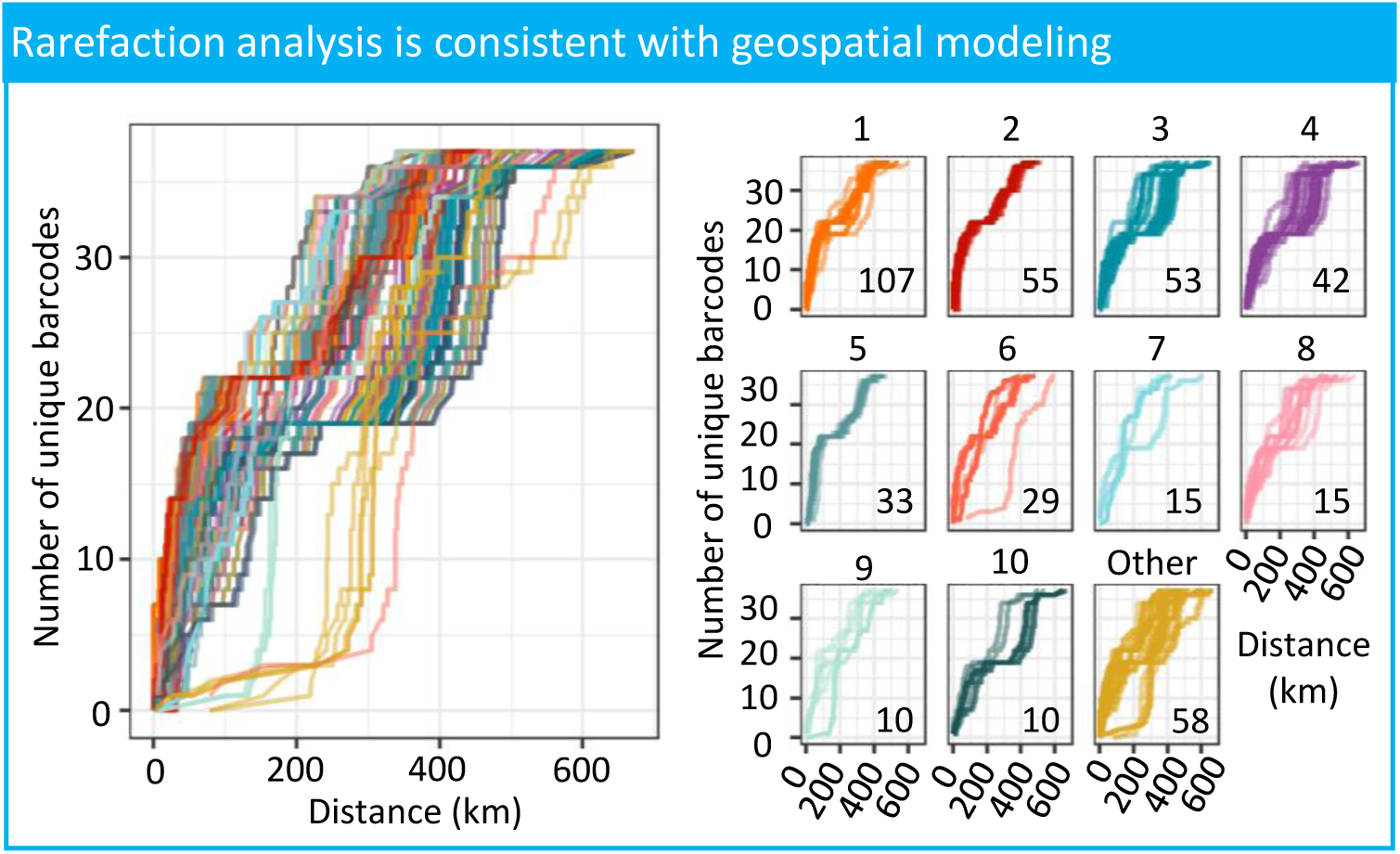
Opportunistic sampling does not drive spatial connectedness of samples. Rarefaction of index worm cases on geographical proximity of barcode diversity shows a consistent addition of diversity in quantitatively identified transmission ranges of barcodes sets (<50 kilometers). The left figure shows all rarefaction analyses for each worm case, and the right figure is separated by the barcode set number (1-10 for common barcodes, other for barcodes found in less than 10 samples). The numbers in the lower right corner are the number of worms in each barcode set.

### 1.10 Expanding genetic markers can improve sensitivity for comparing worm populations

Including mitochondrial variants outside of CO3, cytB, and ND3-5 loci increased the discrimination between worm pairs in an independent population (Fig 4A). Variants were called across the mitochondrial genome from publicly available data for 19 worms from Chad [18]. Forty-four variants were found within the loci sampled in this study, and 132 variants outside of the samples loci range. A shift in the genetic similarity distribution in any direction suggests the new loci can alter the genetic relatedness of a sampled population. We observed a shift in the probability function mass from 0.55 similarity for within loci variants to a second mass at 0.80 for outside loci variants. The distribution of genetic similarity between worms using the 44 loci variants is statistically different than the distribution of genetic similarity using the 132 non-loci variants (Kolmogorov–Smirnov test, p < 0.001). The broader pairwise similarity distribution of outside loci is robust to the number of variants in the pairwise calculation (Fig 4B). Distributions of randomly subsampling 44 non-loci variants compared to the distribution of 44 loci variants showed that in some instances the population was less genetically identical with peaks shifted to the left, and maintaining a higher density of similarity in the 0.75-0.85 range. However, because of the small sample size without accompanying GPS coordinates of this independent population, we cannot extrapolate the effect of barcode sets on the spatial links between identical and non-identical pairs.

**Figure 4:**
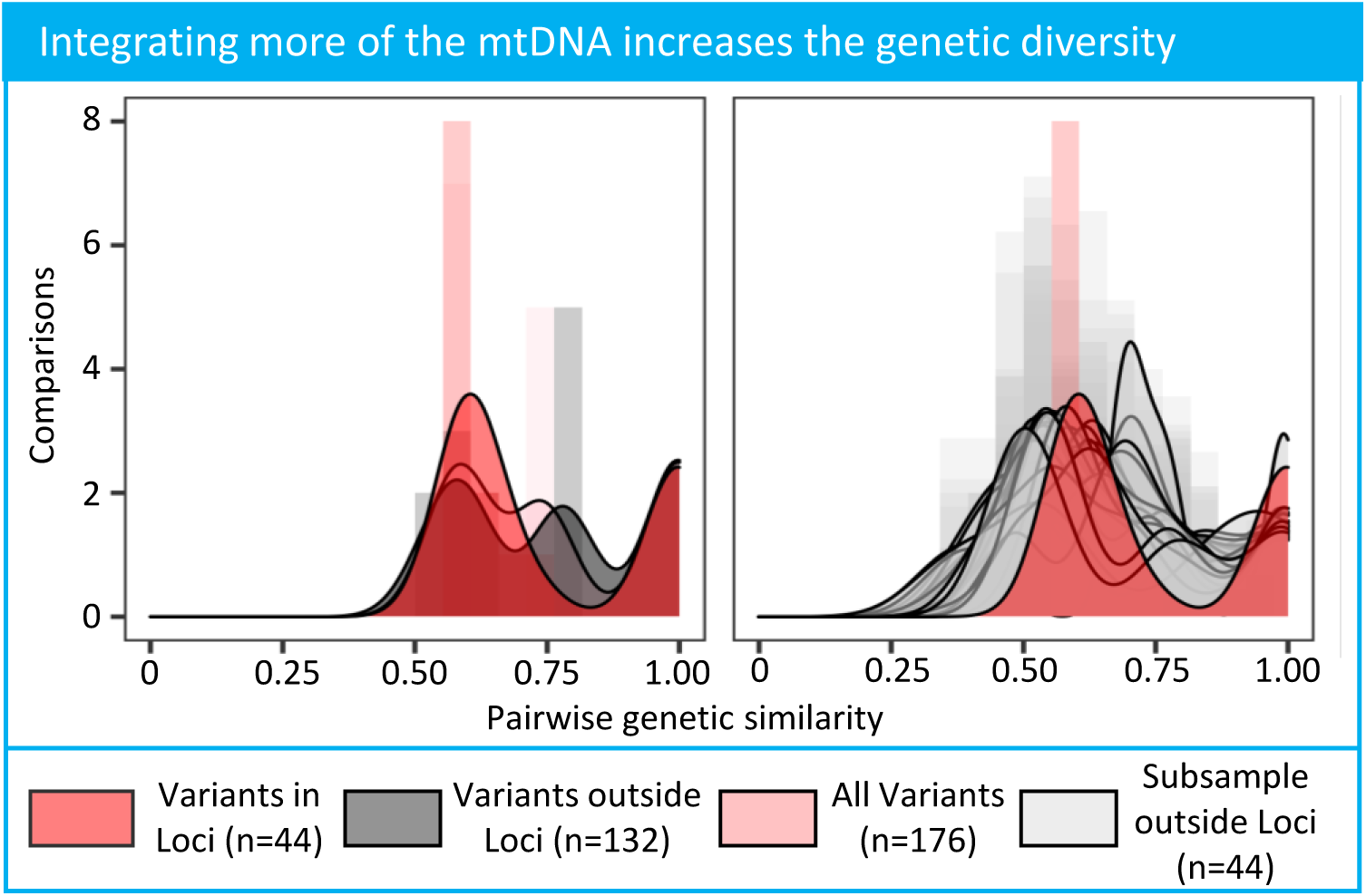
Distribution of all-by-all pairwise genetic similarity of variants within or outside of currently sampled loci. Within loci variants were determined with CO3, cytB, and ND3-5 genomic ranges provided by PlasmoDB. Smoothed density kernel distributions are overlayed on pairwise genetic similairty distribution histograms for the different genomic regions.

Excluding variants found in only one worm, we drop from 176 variants to 134 variants for comparison across the mitochondrial genome. The distribution of outside loci genetic similarity is still broadened in relation to within loci, but the effect is less pronounced (S8 Fig, Kolmogorov–Smirnov test p = 0.001). Distributions of randomly subsampled 34 non-loci variants compared to the distribution of 34 loci variants had a less pronounced decrease in sequence similarity compared to the inclusion of singletons, but confirmed a higher density of similarity in the 0.75-0.85 range. Variants were called with an orthogonal method to circumvent any issues with genotype calling on a small sample size in GATK. More variants across the mitochondrial genome are identified calling with bcftools than GATK (185 versus 176 variants). Variants calls with bcftools replicated different distributions for variants within loci and outside loci with a secondary peak around 0.75 (Kolmogorov–Smirnov test, p = 0.001, S9 Fig).

## Discussion

To our knowledge, this is the largest retrospective study linking surveillance and genetic data to understand Guinea worm transmission in Chad to date, encompassing samples from 2012-2018, excluding 2014. Knowing the range of transmission for genetically linked samples has the potential to be an important monitoring and evaluation tool for the campaign. We identified 41 unique molecular barcodes from the 459 Chadian worm sequences. Among the 459 samples, 426 samples had associated GPS data. The analyses in this study provide quantitative bounds on the geographic range of most transmission within a median 18.6 kilometer radius, and often within an approximately 50 kilometer radius. These results suggest the most effective interventions should consider case sweeps, water monitoring, and abatement approximately 20 kilometers around a reported case to reduce the spread of genetically related parasites. The population distribution falling under 50 kilometers implies that extending these efforts from 20-50 kilometers would largely dampen the dispersion of genetically related worms, with diminishing returns for efforts greater than 50 kilometers from a reported case. However, worms linked by barcode identity are not geographically isolated populations. Each sample is approximately surrounded by worms representing five to ten different barcodes within a 50 kilometer radius (Fig 3), and individual barcodes can vary in their transmission range from three to 150 kilometers. It is important to consider the genetic diversity of worms in an area when guiding geographically imposed interventions from these results.

The modeling results presented in this article are consistent with previous analyses, but also expand the scope of geographic and genetic relatedness. The phylogenetic analysis of barcodes created from all 459 samples in this work supports the earlier conclusion by Thiele et al. [47] that humans and dogs share a similar Guinea worm population in their analysis of 75 samples (S1 Fig). There is not a clear temporal trend between host species and suggests fluid transmission of *D. medinensis* between humans and dogs. Previous genetic analyses using spatial principal components analysis had identified a geographic trend of genetic relatedness down the Chari River in Chad [47, 20, 25]. The research in this article has expanded the scope of that analysis by revealing that genetically identical and near-identical samples cluster geographically for multiple areas across Chad. (Fig 1C).

The findings of this work are also aligned with the known epidemiology of the disease and biology of the parasite. Recent surveillance efforts involving the collaring and GPS tracking of dogs in Chad show dogs visit water sources within a 10 kilometers range, with variation across study sites [36]. In our study, the substantial mass of the genetic similarity distributions around 18.6 kilometers usually within 50 kilometers are broadly in agreement with the dog roaming range including the observed variation by geography; see Fig 2B for reference. Both of these research efforts support the current epidemiological intuition and hypotheses about geographic connectivity of infections and the role of dogs as a reservoir [21, 47, 36]. These analyses provide quantitative bounds on the geographic range of local transmission.

Identifying numerous sets of identical or nearly identical genetic sequences among the 459 samples is also expected due to the parasite biology. Direct maternal mitochondrial inheritance ensures genetic markers on this region are informative of lineage, especially considering the small estimated spontaneous mutation rate or chance of a sequencing error (§1.7). While identifying identical or nearly identical molecular barcodes does not allow us to directly infer transmission events or ancestry, the persistence of barcodes across available years, coupled with the parasite biology, suggests a sustained population of worms related through transmission. Even though the probability of having observed a direct transmission event in the available genetic samples is quite low (given the total number of reported cases), the molecular barcodes demonstrate distinct value even though the transmission network is only partially observed. Taken together, these findings strongly suggest an epidemiological connection identified using the genetic data and can help inform the local epidemiology of Guinea worm in Chad.

There are several limitations to the analysis and modeling in this article. The samples that were sequenced and included in this analysis, specifically for worms collected from dog hosts, were retrospectively selected in order to span the geography of Chad and not through a systematic sampling frame. As mentioned, this lead to an absence of 2014 samples in this study. We have mitigated the challenges posed by constraints on the data and methodologies by evaluating the robustness of each conclusion through a series of sensitivity analyses (Fig 1 and 2). We demonstrated the geographic clustering results are not sensitive to the fact that many of the hosts were multiply infected with Guinea worm which could have artificially inflated the geographic proximity between sample pairs (S5 Fig).

In addition, the characteristics of the *Dracunculus medinensis* genome is not as well-understood as other viruses, bacteria, or parasites. There does not exist a set of tailored phylogenetic or phylodynamic methodologies for Guinea worm parasites [47, 13]. With either a comprehensive baseline genetic diversity from Chad, a known mutation rate for Guinea worm, or both, we could more accurately assess the genetic similarity threshold to cluster worms based on the mutation biology of the parasite. Despite these constraints, we showed that allowing a single base pair difference between worm pair barcodes maintained spatial clustering of identical and nearly identical barcodes relative to unrelated barcodes that have two or more mutations between pairs in the population (S2 Fig). However, a more resolved characterization between pairs of samples or barcodes is potentially limited by the use of three mitochondrial loci coupled with the small sample size relative to the number of observed and unobserved total cases from 2015-2017. To test this hypothesis, we investigated an independent population of genetic samples that had more complete coverage of the mitochondrial genome (§1.10). We were able to identify that more variants are observed in other regions of the mitochondrial genome (§1.10), which was confirmed with a combination of bootstrapping and variant callers. Due to small sample sizes of available whole mtDNA, we were unable to conclude whether the extra variants improve the genetic differentiation within barcode sets and refine the geographic clustering. The substantial number of additional variants in the mitochondrial genome strongly suggests that access to more of the genome will allow better between resolution of genetic similarity for Chadian Guinea worm.

Despite these limitations, the modeling and analyses in this article have important implications for policy makers and elimination programs. From 2010-2019, there has been a concurrent increase in both reported cases and surveillance efforts; a complete characterization of the genetic diversity could help distinguish whether Guinea worm prevalence is actually increasing or is a consequence of improved surveillance. Although there is uncertainty of how genomic signatures are impacted by interventions and the potential temporal lags caused by the year-long life cycle, the continued appearance of genetically identical worms across years suggests genomic data is informative for understanding transmission, surveillance, and even interventions. Monitoring the genetic landscape could provide programmatic evidence for the effectiveness of geographically localized interventions by observing the elimination of barcode lineages. Sustained barcodes are particularly useful in instances where case reports may be disrupted due to insecurity or inaccessibility.

A comprehensive bank of all genomic samples paired with geographic data would allow a broader set of analyses to help the elimination program for outbreak analysis, importation versus local circulation characterization, and potentially reveal unknown animal reservoirs. With the available genetic data, we cannot provide programmatic guidance on a specific number of samples that should be collected and sequenced to capture the genetic diversity in Chad due to the high diversity of the *currently* sampled population (S1 Appendix). Sequencing more historical samples could either decrease genetic diversity and orphaned pairs or support the high diversity of the population. The former would suggest a subset of samples could be sequenced to characterize Guinea worm diversity, while the latter would require sequencing a majority or all of the identified worms. Given current surveillance protocols, samples of the parasite are already collected in addition to a standardized survey for all cases. Thus, sequencing a subset or all of the available retrospective and prospective Guinea worm samples is feasible with respect to the availability of genetic material, an established sequencing protocol, and access to efficient high-throughput sequencing technologies. The confirmed improvement in genetic resolution across the whole mitochondrial genome suggests switching sequencing technologies for the proposed scale up of genetic sequencing of worms is an important future research direction. More broadly, coupling these findings with innovations on constructing phylogenies from whole genome sequencing of the Guinea worm larvae [13] and microsatellites of the worm nuclear genome [47], a holistic program of sequencing technologies and analytic methodologies will help translate research insights into programmatic input for the elimination of Guinea worm in Chad. Furthermore, additional analyses of the full range of epidemiological data collected by the CGWEP alongside linked genomic data are warranted, and may further elucidate transmission dynamics in Chad. The parasite genome has the potential to be an integral tool for the end-game strategy in Chad and beyond.

## Data Availability

Genomic data and source code will be made available upon publication. Requests for epidemiological data that includes case coordinates must be submitted to and approved by the Chad Guinea Worm Eradication Program.

## Acknowledgments

We would like to thank Dr. Fernando Torres for his helpful discussions regarding Guinea worm eradication efforts and Dr. Albert Lee for modeling discussions. This work would not be possible without the field and surveillance teams of the national Guinea worm eradication program in Chad. Thank you for the efforts of all involved to collect the epidemiological data and samples.

## Disclaimer

The findings and conclusions in this report are those of the author(s) and do not necessarily represent the official position of the Centers for Disease Control and Prevention/the Agency for Toxic Substances and Disease Registry.

## Supporting information

**S1 Table:**
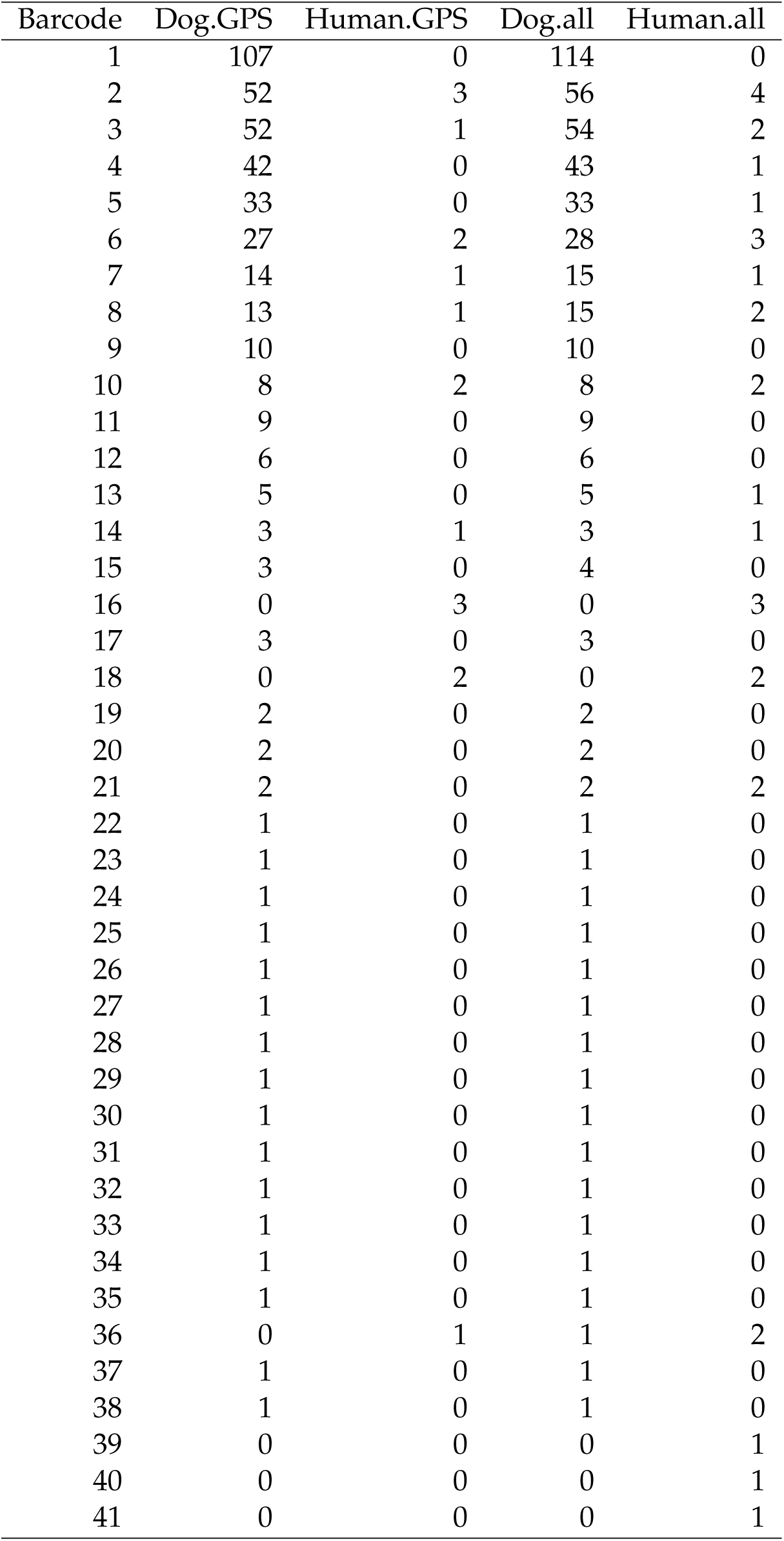
Barcode counts for sequenced samples. GPS columns indicates the number of samples for each barcode used in geospatial analyses by species. All worms indicates the number of worms assigned to each barcode, including samples that were not linked to GPS data for each species. Phylogenetic relationships for barcodes (see S1 Fig) are plotted once per species, but have different numbers of samples represented in each branch.

| Barcode | Dog.GPS | Human.GPS | Dog.all | Human.all |
| --- | --- | --- | --- | --- |
| 1 | 107 | 0 | 114 | 0 |
| 2 | 52 | 3 | 56 | 4 |
| 3 | 52 | 1 | 54 | 2 |
| 4 | 42 | 0 | 43 | 1 |
| 5 | 33 | 0 | 33 | 1 |
| 6 | 27 | 2 | 28 | 3 |
| 7 | 14 | 1 | 15 | 1 |
| 8 | 13 | 1 | 15 | 2 |
| 9 | 10 | 0 | 10 | 0 |
| 10 | 8 | 2 | 8 | 2 |
| 11 | 9 | 0 | 9 | 0 |
| 12 | 6 | 0 | 6 | 0 |
| 13 | 5 | 0 | 5 | 1 |
| 14 | 3 | 1 | 3 | 1 |
| 15 | 3 | 0 | 4 | 0 |
| 16 | 0 | 3 | 0 | 3 |
| 17 | 3 | 0 | 3 | 0 |
| 18 | 0 | 2 | 0 | 2 |
| 19 | 2 | 0 | 2 | 0 |
| 20 | 2 | 0 | 2 | 0 |
| 21 | 2 | 0 | 2 | 2 |
| 22 | 1 | 0 | 1 | 0 |
| 23 | 1 | 0 | 1 | 0 |
| 24 | 1 | 0 | 1 | 0 |
| 25 | 1 | 0 | 1 | 0 |
| 26 | 1 | 0 | 1 | 0 |
| 27 | 1 | 0 | 1 | 0 |
| 28 | 1 | 0 | 1 | 0 |
| 29 | 1 | 0 | 1 | 0 |
| 30 | 1 | 0 | 1 | 0 |
| 31 | 1 | 0 | 1 | 0 |
| 32 | 1 | 0 | 1 | 0 |
| 33 | 1 | 0 | 1 | 0 |
| 34 | 1 | 0 | 1 | 0 |
| 35 | 1 | 0 | 1 | 0 |
| 36 | 0 | 1 | 1 | 2 |
| 37 | 1 | 0 | 1 | 0 |
| 38 | 1 | 0 | 1 | 0 |
| 39 | 0 | 0 | 0 | 1 |
| 40 | 0 | 0 | 0 | 1 |
| 41 | 0 | 0 | 0 | 1 |

**S1 Fig:**
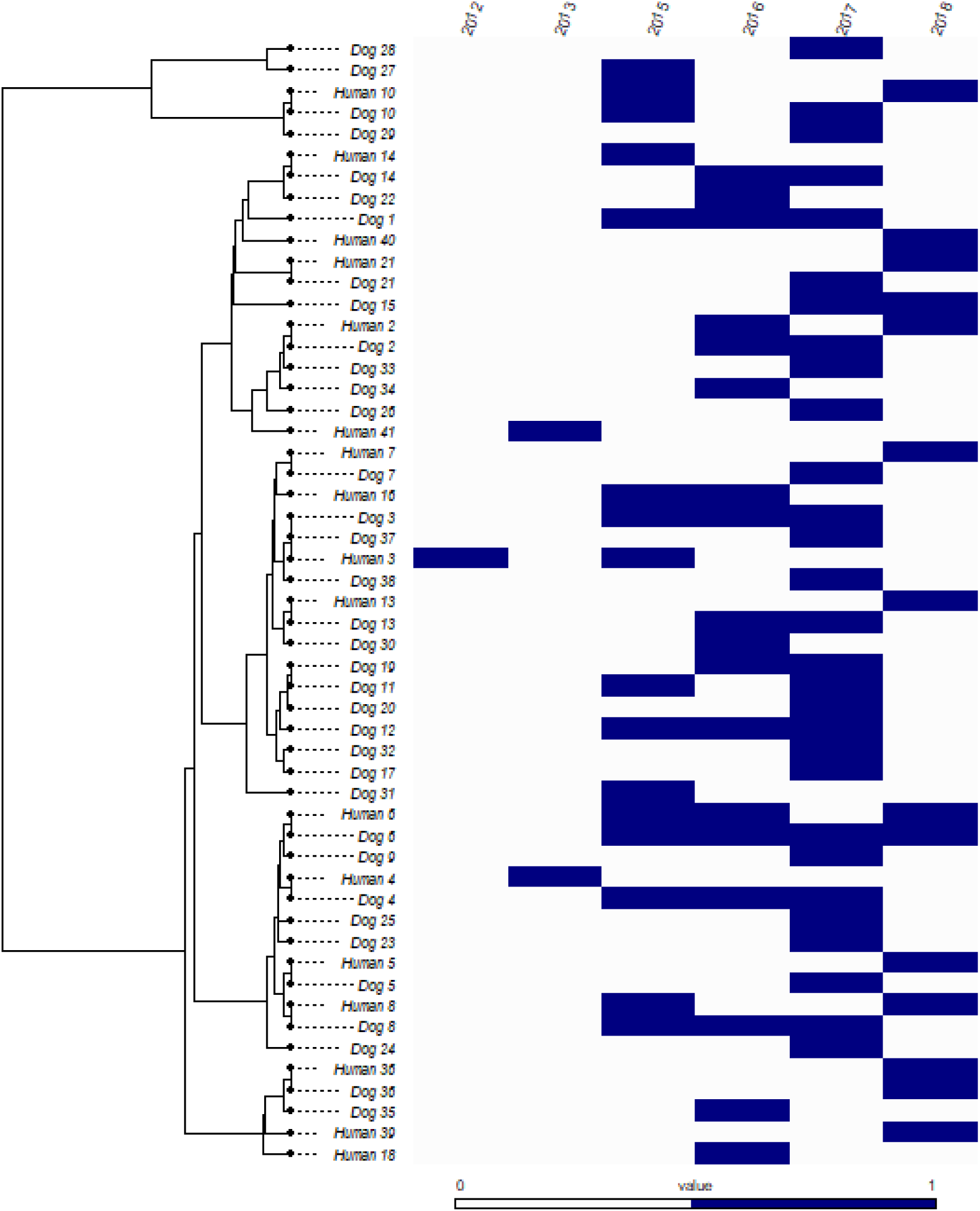
Phylogenetic comparison of worm barcodes identified per species. This graph demonstrates that barcodes were shared among humans and dogs alike, and were often present over the course of multiple years. Tree tip labels represent a single barcode per species. Refer to S1 Table for sample counts of each barcode by species. Barcode identifying numbers are represented more than once if observed in both species. A blue filled cell next to each barcode is indicative that barcode was represented at least once in the respective year.

**S2 Fig:**
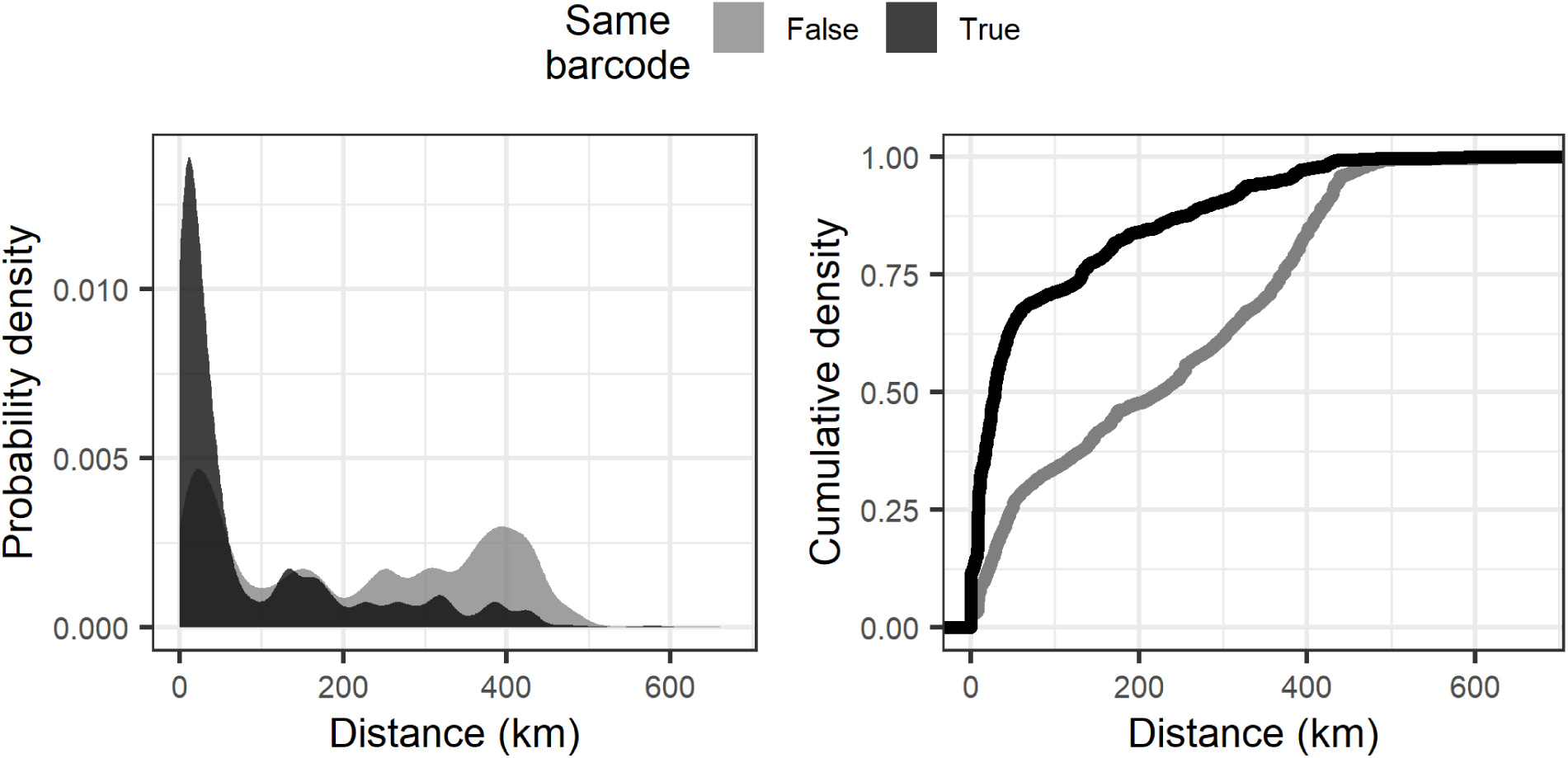
Spatiotemporal modeling of barcode relatedness collapsed by a single base pair difference. Number of pairwise comparisons for identical barcodes = 13, 967, for non-identical barcodes = 76, 558.

**S3 Fig:**
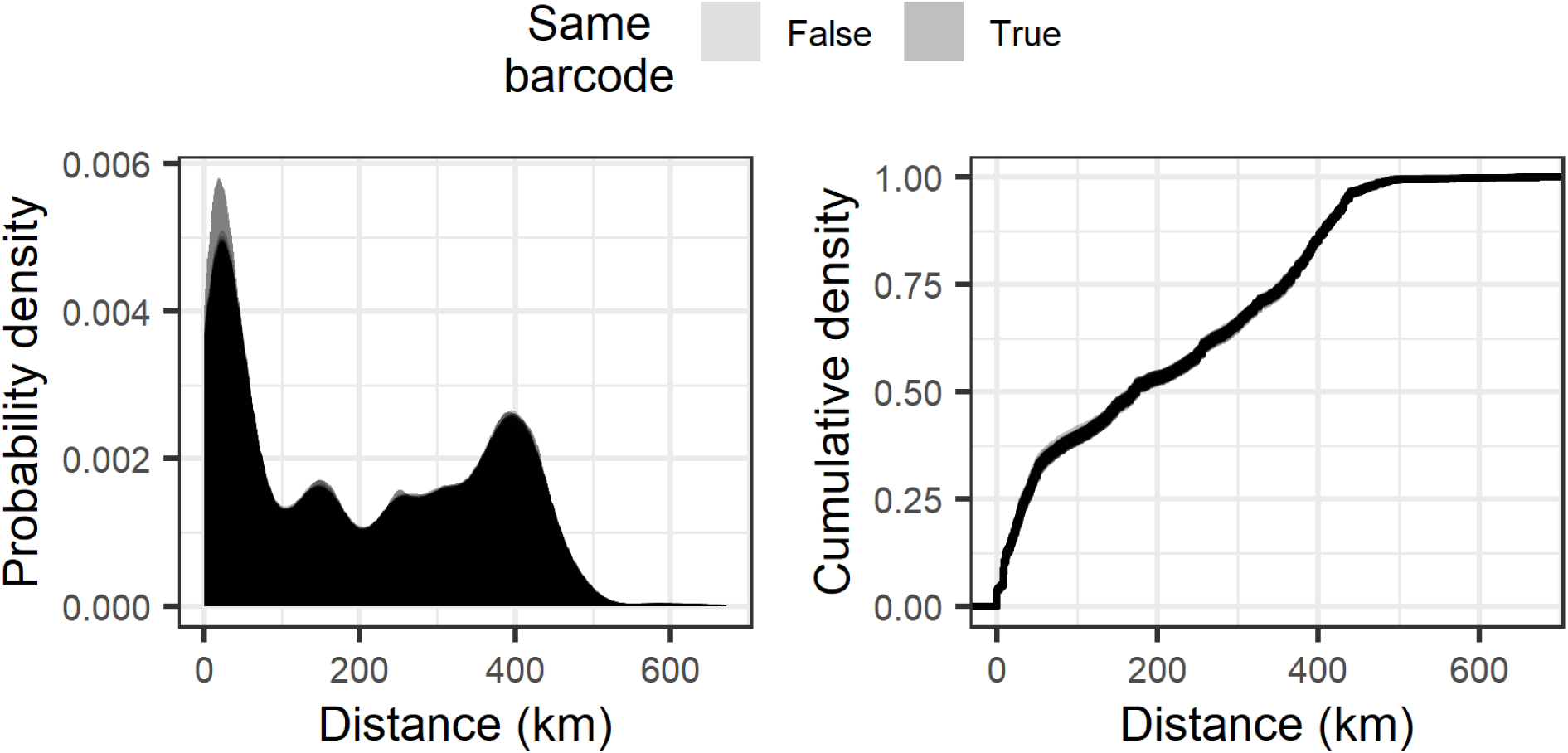
Permutation of distance between cases on barcode relatedness densities. The x-axis is the permuted distance between worm pairs and the y-axis represents the density for population genetic similarity scores for all worms (n=426). Each line represents worm pair distance permutation (n=100) for the population. The lines for identical and non-identical barcodes are consistent between permutations and overlap.

**S4 Fig:**
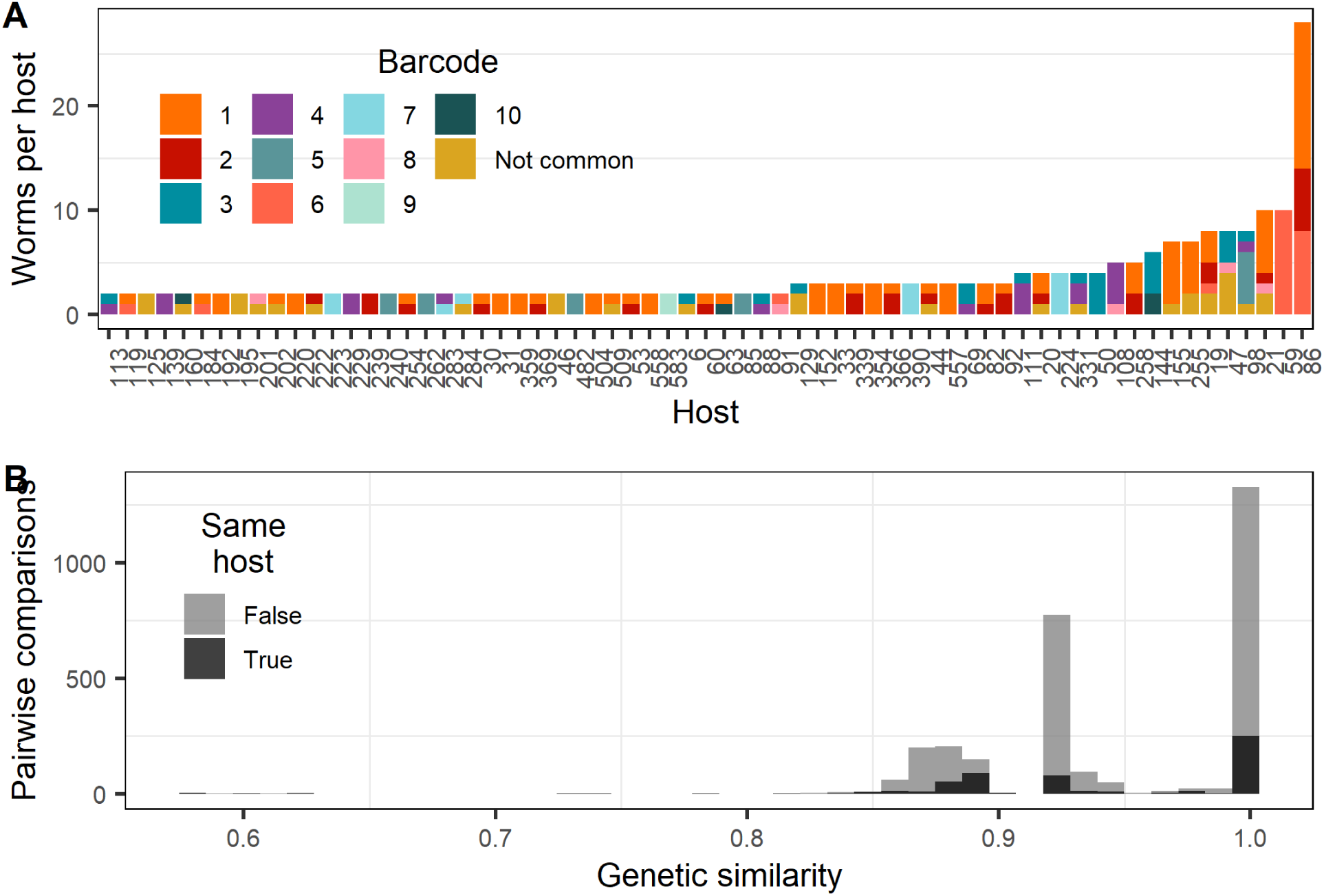
Barcode identity within hosts and genetic similarity distribution by shared location. A. Each host with the number of worms pertaining to each barcode set. “Not common” refers to barcodes found in less than 10 samples in the population for visual clarity. B. Distributions of genetic similarity between worms with the same reported GPS coordinates, colored by whether the pair is obtained from the same or different hosts.

**S5 Fig:**
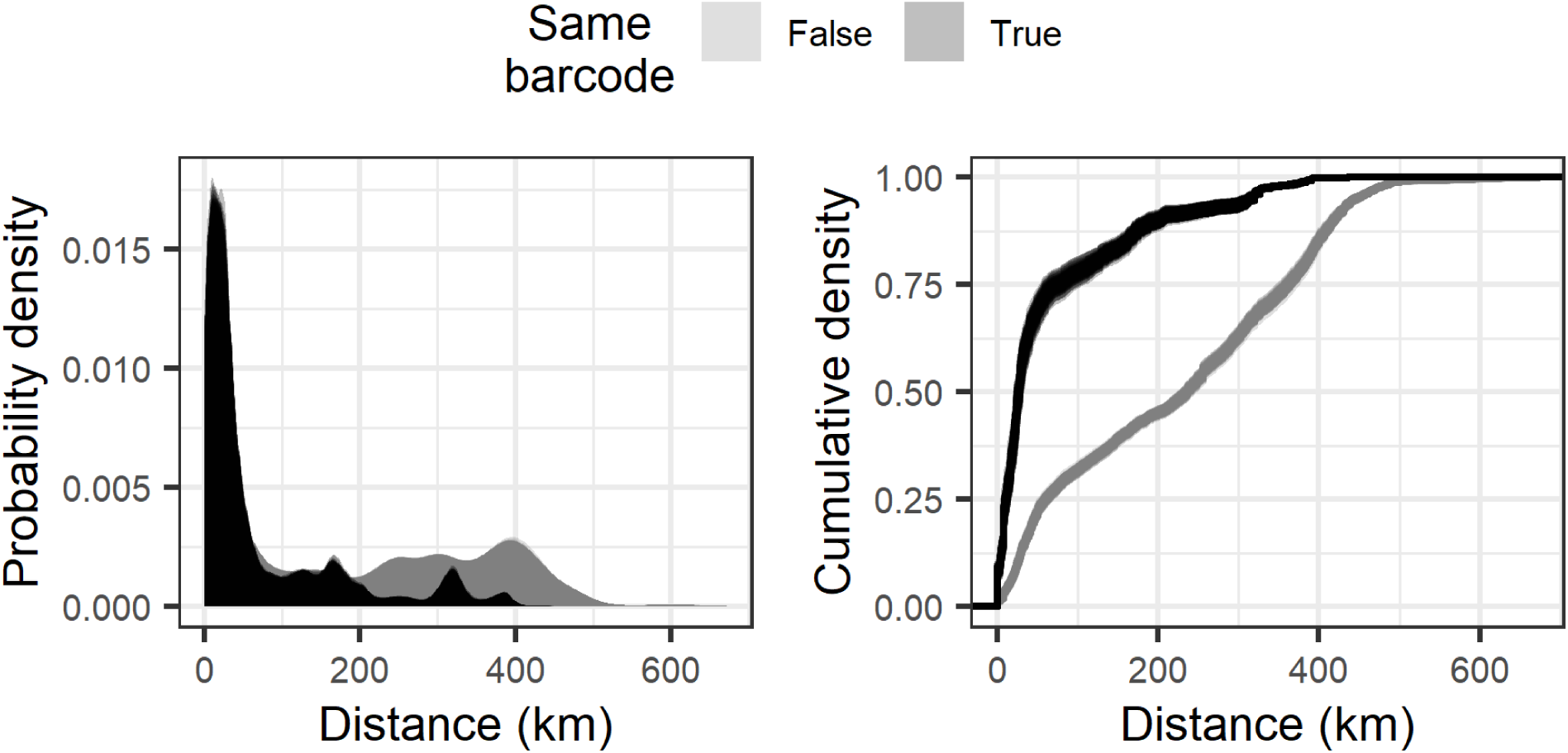
Host subsampling on barcode relatedness densities. The x-axis is the distance between worm pairs and the y-axis represents the density for population genetic similarity scores with one worm per host (n = 282). Each line represents a bootstrap (n=100) of a single worm per host.

**S6 Fig:**
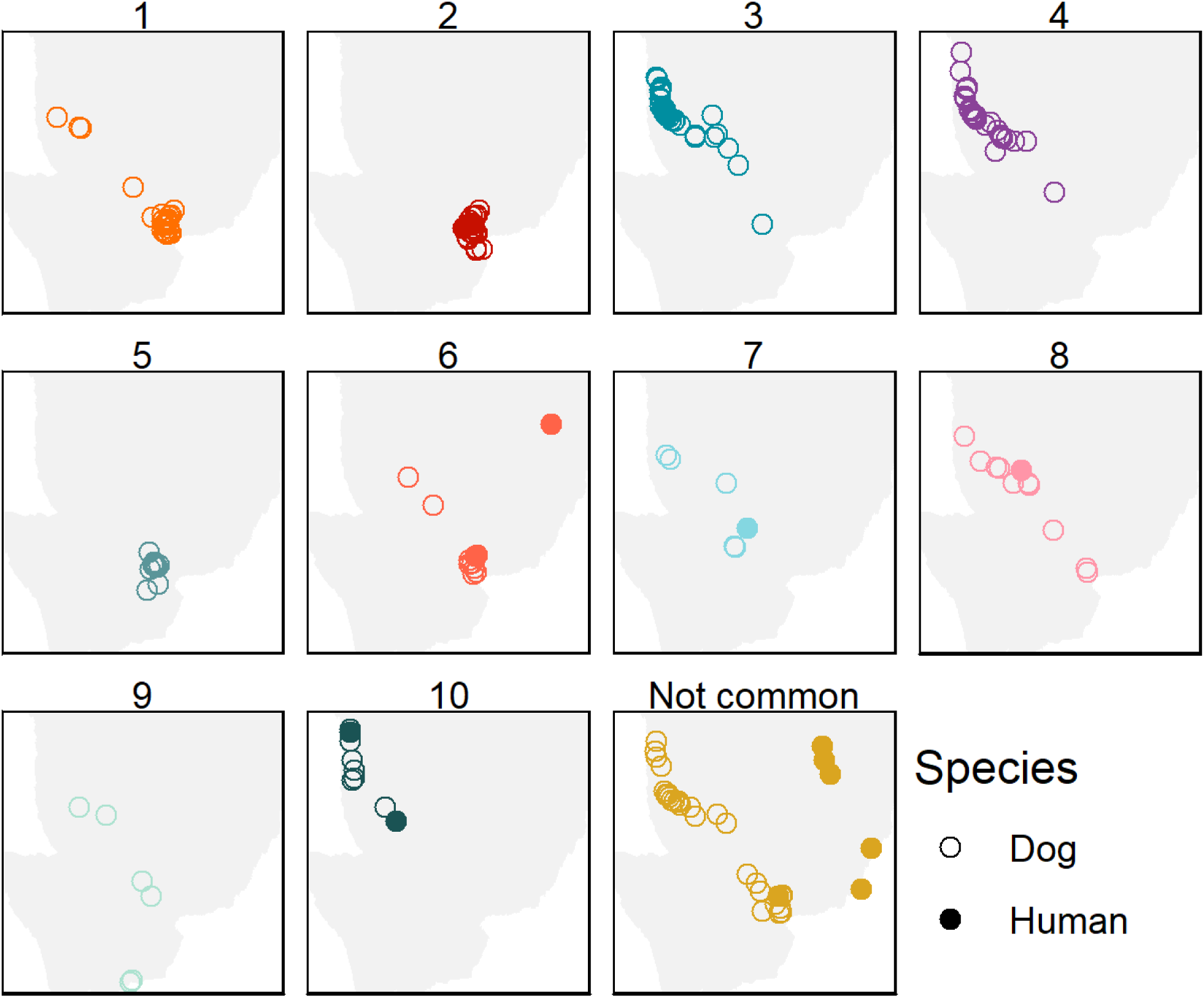
Location of common barcodes in Southern Chad. Samples are colored by their barcode identifier, and shapes represent the host species. Barcodes 3,4, and 10 appear have similiar spatial distributions. Barcodes 2 and 5 have similar spatial distributions and are highly focal. some barcodes span a very large geographic area, which could suggest they are ancestral sequences that have diffused over time or are transmitting due to human behaviors.

**S7 Fig:**
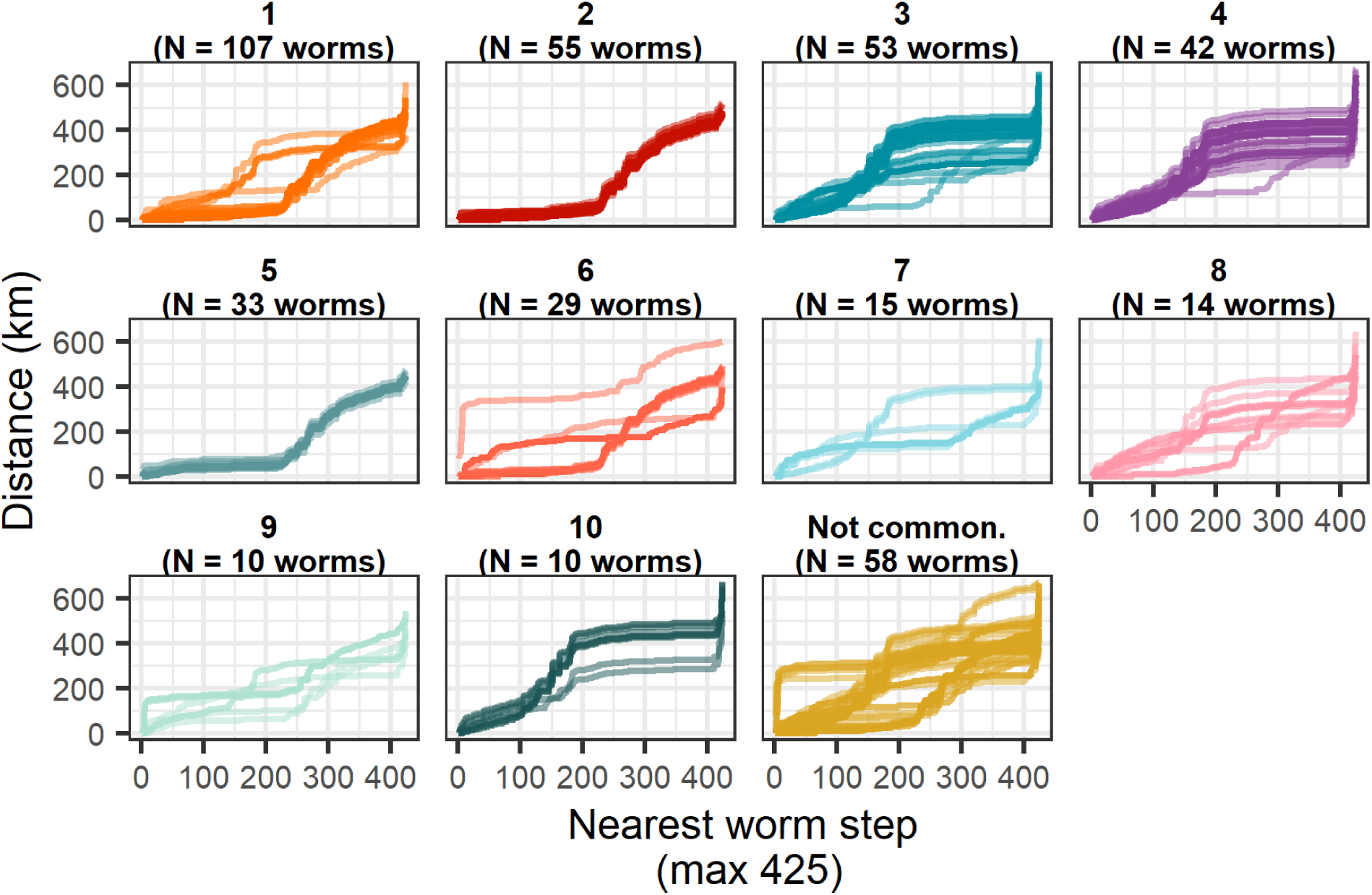
Geographic distance between cases for each worm. The geographic distance between worms relative to a single worm were organized in ascending order. The x axis is the worm index by distance and the y axis is the cumulative distance from each index worm. Spans of a flattened curve are indicative of geographic stretches that do not contain any samples and support the lack of barcode diversity observed in cerain geographic distances in Fig 3.

**S8 Fig:**
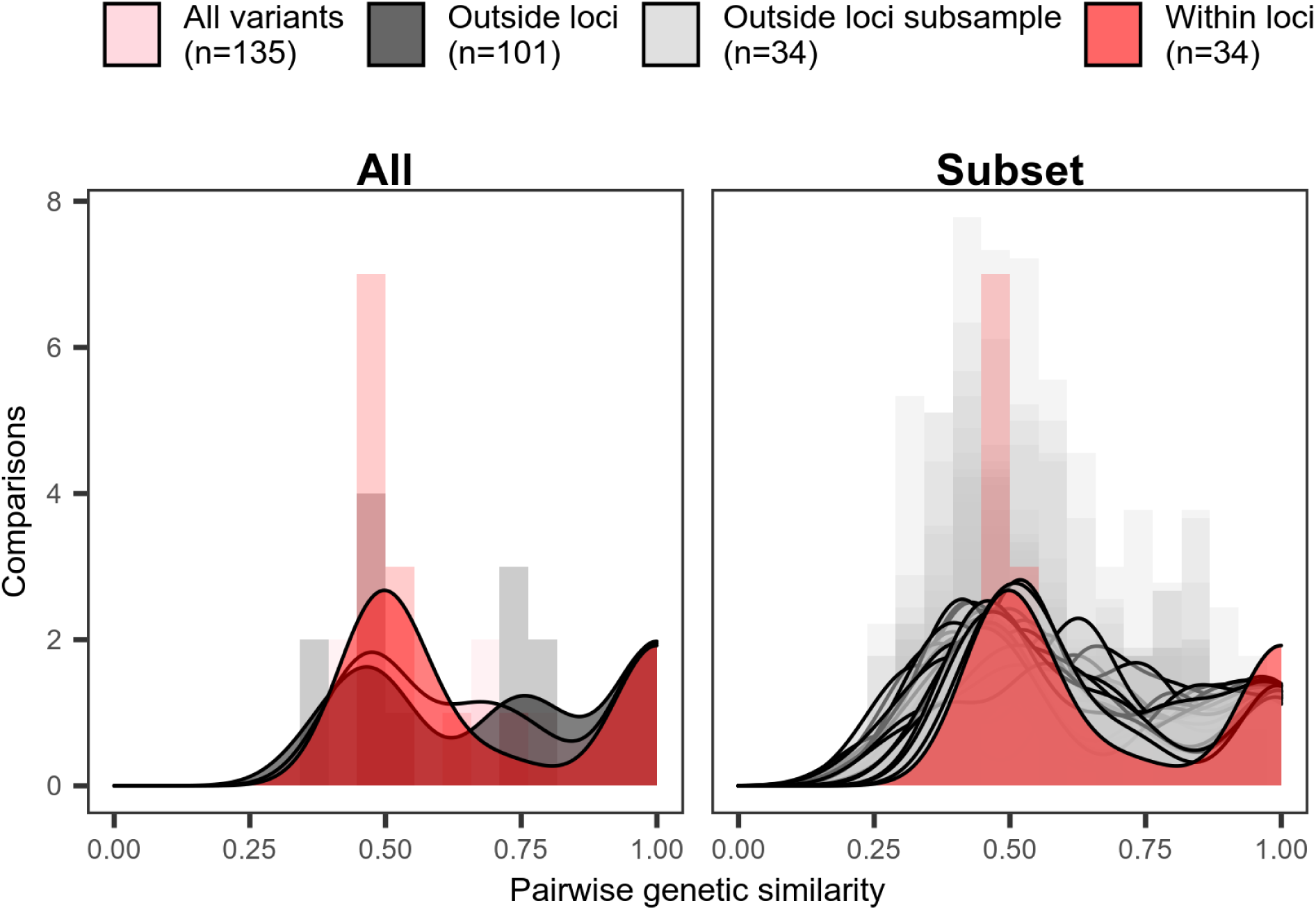
Distribution of pairwise similarity using GATK variants found in greater than one worm. The x-axis is the measured genetic similarity for variants within current loci (n=34) and extending to the rest of the mitochondria (n=101), and y-axis is the number of pairwise comparisons (19*19 = 361). Filled regions show the smoothed density estimates for histograms.

**S9 Fig:**
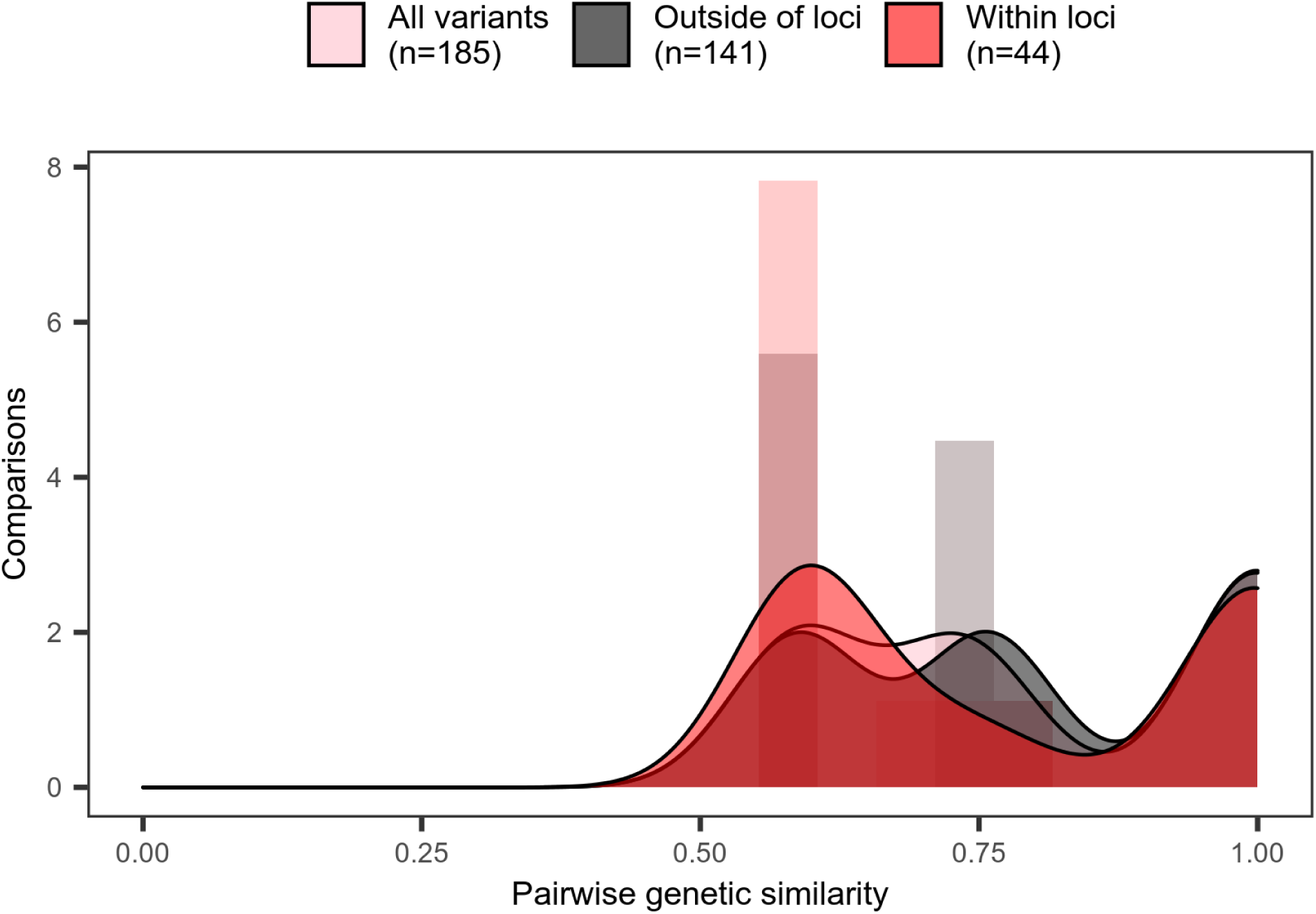
Orthogonal validation of population pairwise similarity using variants identified by bcftools mpileup. The x-axis is the measured genetic similarity for variants within current loci (n=44) and extending to the rest of the mitochondria (n=141), and y-axis is the number of pairwise comparisons (19*19 = 361). Filled regions show the smoothed density estimates for histograms.

