## Supplementary material for "Linked surveillance and genetic data uncovers programmatically relevant geographic scale of Guinea worm transmission in Chad": S1 Appendix

### 1 Predicting the increase in barcode variety with additional sequencing

We observed 41 unique barcodes from sequencing 459 genomes. 459 is a subset of the number of available samples and further sequencing will intuitively identify previously unobserved new barcodes. The result of this analysis aims to provide a quantitative assessment and guidance for the Guinea worm program for how many samples are required to recover the underlying genetic diversity.

#### 1.1 Fitting a negative binomial distribution to unique barcodes

A simple attempt to extrapolate the number of unique barcodes that may be uncovered from the current data, we fit a negative binomial distribution up to 5000 samples that followed the distribution of unique barcodes in the observed samples. In this process, barcodes are represented by a (unspecified) natural number and sampling from a distribution over natural numbers will yield more unique elements over time.

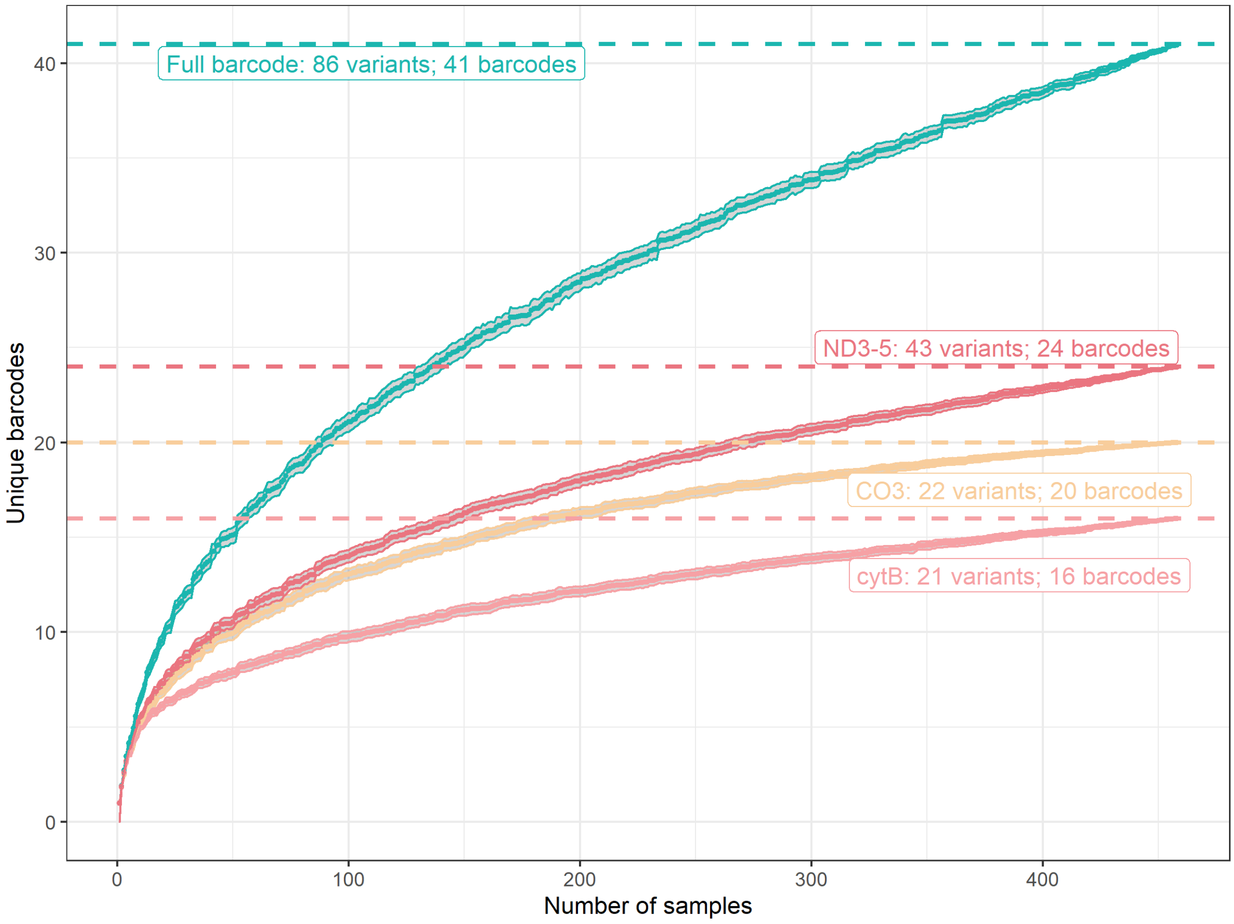


Figure 1: Accumulation curve of 459 samples barcodes, with 90% prediction intervals derived from 100 replicates.

Given such a distribution, the mean number observed barcodes $N_o$ after $m$ samples is

$$E(N_o|m,\theta) = \sum_{x=0}^{\infty}(1-(1-p(x))^m),$$

where p(x) is the mass function of the negative binomial distribution and $\theta$ its parameters. The distribution for unique barcodes for each subsample size up to 459 was counted for 100 replicates. The mean unique barcodes for each subsample size is plotted with a 90% confidence interval from the replicates.

To determine the Negative Binomial parameters appropriate to extrapolate up to 5000 samples, we replicated the observed population diversity curve using the mean of 100 negative binomial replicates with a dispersion parameters ranging from 0.1 to 0.3 with combinations of success probability for 0.025 to 0.050. The negative binomial line which best aligned to the saturation curve of the 459 samples was visually chosen, with dispersion parameter 0.20 and success probability of 0.039. The number of successes in the negative binomial draws were used as a barcode identifier for uniqueness

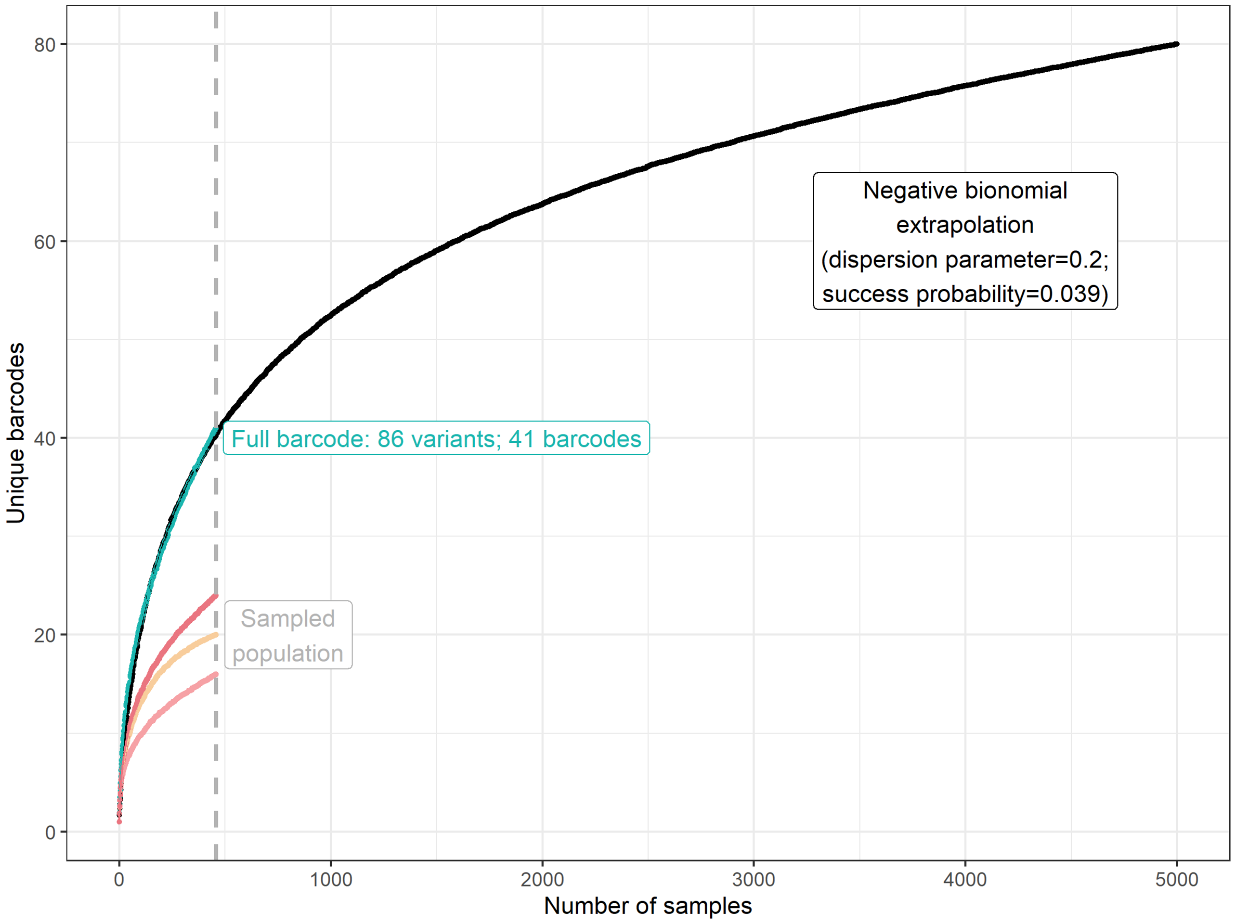


Figure 2: Extrapolation of the Negative Binomial up to 5000 samples.

of subsamples up to 5000.

This method is limited by not retaining the identity of barcodes through the process. While the distribution of sampled barcode counts resembles a negative binomial distribution, the tail of the distribution is not uniform; a few barcodes are observed in a high number of samples, but there is a large gap between barcodes seen in a few hundered samples and observed less than 10 times in the sampled population. The negative binomial as modeled does not account for count gaps between the low and high frequency barcodes, assuming equal sampling probability in this range and not a higher likelihood of sampling from a more frequent barcode in the population. Hence, an extended empirical Bayes approach was explored in further depth.

### 1.2 Empirical Bayes

Let $N$ be a hypothetical (super-)population of barcodes. A combinatorial limit exists for the number of possible barcodes based on its length and perhaps other features, though in practice it is so large that we ignore if for now. We take $N$ not to be the size of such a space, but still a large number of distinct barcodes in the population. We will not consider phylogenies as part of the analysis; each barcode will be considered a separate, unrelated species.

Let $M$ be a large population of samples, for which sequencing all is unlikely. $M$ may represent the total known population. We may allow $N$ to be much greater than $M$; that is, we allow a superpopulation of barcodes. Similarly, let $n$ be the number of unique barcodes observed in a sequenced sub-sample of size $m \leq M$, assumed to be drawn at random.

With $\mathbf{p}$ the $N$-dimensional parameter of relative abundance and $\mathbf{Y}$ the number of each barcode drawn from the $m$ samples. Let

$$\begin{aligned}\mathbf{Y}|\mathbf{p} &\sim \text{Multinomial}(m, \mathbf{p}) \\ \mathbf{p} &\sim \text{Dirichlet}(N, \alpha\mathbf{1_N}).\end{aligned}$$

The prior for $\mathbf{p}$ is governed by a single parameter $\alpha$, assuming that $N$ is known. This implies no prior belief of higher relative abundance of specific barcodes. Note that $\alpha = 1$ implies a uniform distribution on the $N$-dimensional simplex. As $\alpha$ increases, the mass of $\mathbf{p}$ becomes more concentrated around equal $p_i = 1/N$; as $\alpha$ decreases, mass become more "polar" (towards 0 and 1), implying that some barcodes are expected to dominate in terms of relative abundance while not knowing which ones they are.

This induces a Dirichlet-Multinomial marginal model, the multivariate analogue to the Beta-Binomial distribution formed by a Binomial distribution with a Beta-distributed $p$. The Dirichlet distribution is a conjugate prior for the Multinomial distribution, with

$$\mathbf{p}|\mathbf{y}, \alpha, N, m \sim \text{Dirichlet}(\alpha_i = \alpha + y_i)$$

This is a useful property, as then the posterior predictive distribution for future samples is Dirichlet-Multinomial with the updated parameters (conditional on known $\alpha$).

An immediate use of this property is the expected number of new barcode detections with further sequencing. Given $p_i$, the probability that barcode $i$ is unobserved in a sample of size $m$ is $(1-p_i)^m$. Thus, the expected number of observed barcodes given $\mathbf{p}$ is

$$\begin{aligned}E(N_o|m, \mathbf{p}) &= E(\sum_{i=1}^{N} 1_{[Y_i>0]}|\mathbf{p}) \\ &= N - E(\sum_{i=1}^{N} 1_{[Y_i=0]}|\mathbf{p}) \\ &= N - \sum_{i=1}^{N}(1-p_i)^m.\end{aligned}$$

Marginalizing over $\mathbf{p}$ where $\mathbf{p} \sim \text{Dirichlet}(\boldsymbol{\alpha})$, for the general case of possibly distinct $\alpha_i$ and observing that $p_i \sim \text{Beta}(\alpha_i, \sum_{j \neq i} \alpha_j)$,

$$E(N_o | m, \boldsymbol{\alpha}) = N - \sum_i \frac{B(\alpha_i, \sum_{j \neq i} \alpha_j + m)}{B(\alpha_i, \sum_{j \neq i} \alpha_j)}.$$

As this expression is for general (known) $\boldsymbol{\alpha}$ we may use it to determine the expected new barcode detections for subsequent sampling, given the $n$ observed barcode detections from the first $m$ samples and an update of $\boldsymbol{\alpha}$. Consider the next $m^*$ samples: $n$ have already been observed, narrowing interest on the $N - n$ remaining barcodes not yet observed.

$$\begin{aligned}
E(N_o^* | m^*, \mathbf{y}, \alpha) &= (N - n) - \sum_{i: y_i = 0} E((1-p)^{m^*} | \boldsymbol{\alpha}); \alpha_i = \alpha + y_i \\
&= (N - n) - \sum_{i: y_i = 0} \frac{B(\alpha + y_i, m^* + \sum_{j \neq i} \alpha + y_i)}{B(\alpha + y_i, \sum_{j \neq i} \alpha + y_i))} \\
&= (N - n) - \sum_{i: y_i = 0} \frac{B(\alpha, (N-1)\alpha + m^* + m)}{B(\alpha, (N-1)\alpha + m}; \sum y_i = m \\
&= (N - n) - (N - n) \frac{B(\alpha, (N-1)\alpha + m^* + m)}{B(\alpha, (N-1)\alpha + m},
\end{aligned}$$

with the last step due to $\alpha + y = \alpha$ for the previously unobserved barcodes.

Equipped with these set of methods, we well examine the expected trajectory for $M = 5000$ samples given the detection from the first $m = 459$ samples. Our approach will be to find the maximum likelihood estimate of $\alpha$ given a choice of $N$ from the observed frequencies (0 for $N - n$ of the barcodes). We will then use $\hat{\alpha}$ as a "plug-in" estimate for subsequent updating and for estimating the expected trajectory. To aid in this comparison – and to preview the later part of the analysis – we also produce a mean trajectory and pointwise 95% intervals for how detection might have occurred for the first 459 samples by random permutations of the order for the observed barcodes.

The results in Figure 3 suggest potentially convergent behavior as $N$ increases, and as one would expect there is relationship between $N$ and $\hat{\alpha}$ (not shown). For our purposes, we will choose a large $N$ with which to continue the analysis, choosing $N = 10000$.

We now use the posterior predictive distribution to produce mean and pointwise prediction intervals for the number of new barcodes that might result from sequencing more samples, up to $M = 5000$. Our approach will be to use the Multinomial-Dirichlet distribution, equivalently a draw from $\mathbf{p} | y_i, \hat{\alpha} \sim \text{Dirichlet}(\alpha_i = \hat{\alpha} + y_i)$ followed by a sequence of samples with these probabilities. This simulated sequence is processed to produce the number of detected barcodes as a function of number of samples.

The results are plotted in the Figure 4 and tabulated in Table 1 for a set of 500 simulations.

There are several caveats with this approach. As mentioned earlier, we have not integrated the genetic relationships between observed barcodes in this analysis. Moreover, we have assumed that the 459 samples are plausibly a random sample of a larger set of potential samples and does not include additional structure to the sampling; the existence of explicit or implicit strata could change

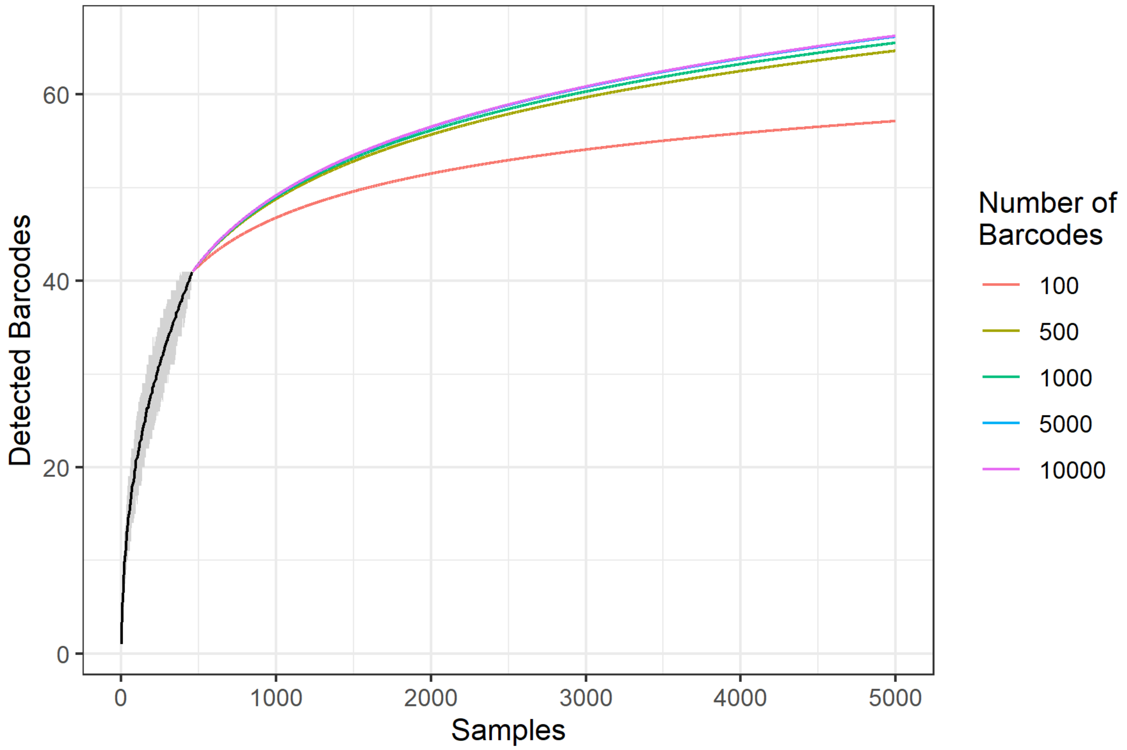


Figure 3: Expected trajectories as a function of $N$ and $\hat{\alpha}$; $\hat{\alpha}$ is a function of $N$ and the observed frequencies.

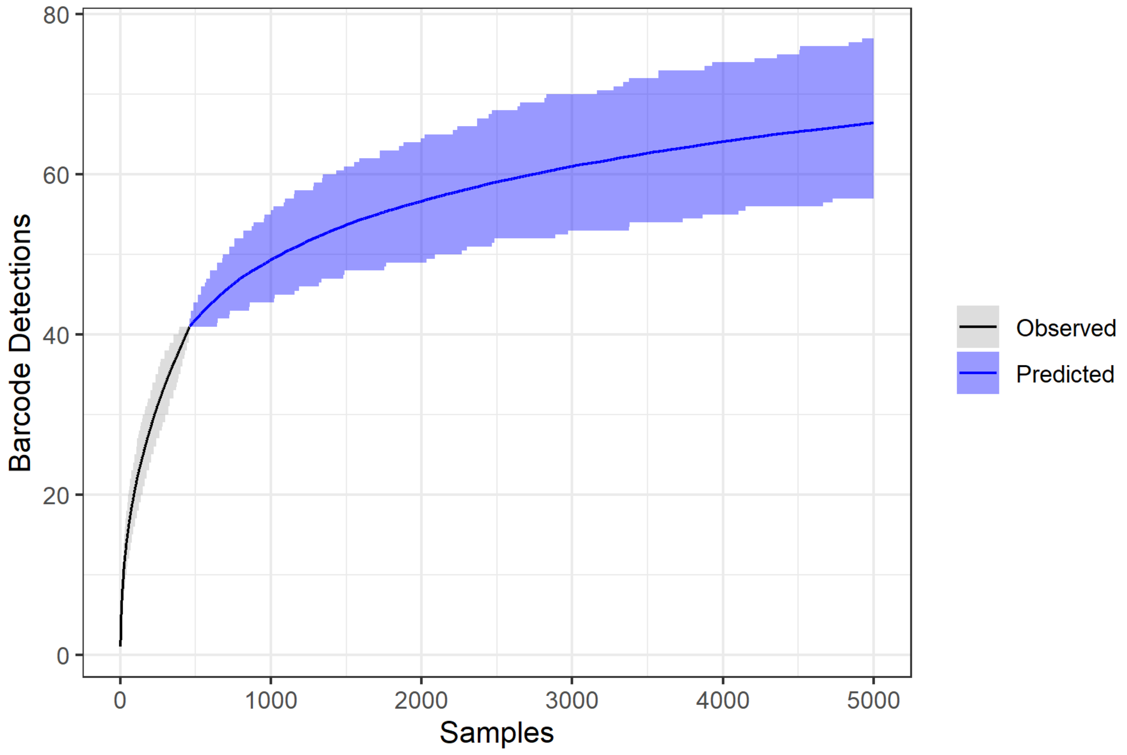


Figure 4: Detected barcodes from 500 simulations, with pointwise 95% prediction intervals.

Table 1: Summary statistics for 500 simulated barcode samples.

| Samples | Mean | StdDev | Min | Max | q0.025 | q0.975 | Epoch |
|---|---|---|---|---|---|---|---|
| 459 | 41.00 | 0.00 | 41 | 41 | 41.00 | 41.00 | Observed |
| 1000 | 49.38 | 2.87 | 43 | 60 | 44.00 | 55.00 | Predicted |
| 2000 | 56.64 | 3.89 | 47 | 71 | 49.00 | 64.52 | Predicted |
| 5000 | 66.42 | 4.95 | 53 | 82 | 57.00 | 77.00 | Predicted |

the model and results due to accounting for between and within stratum variance and the stratified structure of the unobserved barcodes.

Last, we note that this bears a resemblance to a classic treatment of species detection by Fisher et al [2], described in [1]. In that treatment, counts of a butterfly species are modeled as a Negative Binomial distribution arising from a Poisson with Gamma-distributed intensities. Estimation of the parameters of the Negative Binomial distribution can be used to study potential outcomes for known but unobserved species (as well as the observed) in a subsequent study interval.

The connection with our approach arises as follows. If barcodes were detected according to a Poisson distribution with differing intensities $\lambda_i$, then conditioning on a total number of detections $m$, the number of detections of each barcode is $\mathbf{Y}|m, \boldsymbol{\lambda} \sim \text{Multinomial}(m, p_i = \lambda_i / \sum \lambda_j)$. If $\lambda_i \sim \text{Gamma}(\alpha, \beta)$ (iid), then $\lambda_i / \sum_j \lambda_j \sim \text{Dirichlet}(\alpha)$, invariant with respect to $\beta$, and we arrive at our formulation.
